# Inflammatory bowel disease burden in Asia from 1990 to 2019 and predictions to 2040

**DOI:** 10.1101/2023.12.04.23299416

**Authors:** Xingchen Wang, Jiannan Xiong, Zhao Ding, Yueting Liu, Chongguang Yang, Pengfei Xu

**Author notes:** **Corresponding authors:** Department of Hepatobiliary and Pancreatic Surgery, Zhongnan Hospital of Wuhan University, School of Pharmaceutical Sciences, Wuhan University, Wuhan, China, 430072, (PX).

## Abstract

**Background:** While the incidence of inflammatory bowel disease (IBD) has stabilized in Western countries since 1990, it is experiencing an upward trend in newly industrializing countries. The Asian region encompasses a multitude of developing countries at varying stages of IBD progression. Therefore, comprehending the current epidemiological characteristics of the disease in this region becomes imperative, enabling countries and locales in Asia to proactively address the evolving IBD burden in the upcoming several decades.

**Methods:** We analyze variation trends in the burden of IBD in Asia from 1990 to 2019, employing data and methods from the Global Burden of Diseases, Injuries, and Risk Factors Study (GBD), and provide projections for future changes in IBD incidence in Asia over the next 20 years.

**Results:** In 2019, the number of incidence cases of IBD in Asia was 145,561 (95% UI 124,960-170,895), the total number of prevalence cases reached nearly 2 million (95% UI 1.71-2.32), and 13,957 (95% UI 11,898-16,021) patients died of IBD. Meanwhile, the total years lived with disability (YLDs) attributed to IBD amounted to 299,663 (95% UI 198,365-418,635), while the total disability-adjusted life-years (DALYs) rose to 649,760 (95% UI 530,395-783,181). The total number of incidence cases in Asia is projected to reach 179,756 in 2040, with an age-standardized incidence rate of 2.92 per 100,000 population.

**Conclusions:** The increase in the overall burden of IBD in Asia is primarily driven by population growth and aging, with both incidence and DALYs continuing to rise in most countries. It is imperative for each country to adapt its measures to local conditions, improve prevailing healthcare service patterns, and draw insights from the frontier countries to respond to the evolving epidemic characteristics of IBD.

## Background

Inflammatory bowel disease (IBD) is a chronic and potentially life-threatening inflammatory disorder of the gastrointestinal tract, encompassing Crohn’s disease (CD) and ulcerative colitis (UC).^1, 2^ IBD is a life-long condition that typically develops in early life (ages 18-35) in both males and females and is characterized by recurrent episodes of clinical symptoms interspersed with periods of remission.^3, 4^ The principal symptoms of IBD comprise diarrhea, abdominal pain, and rectal bleeding. Additionally, it can involve the joints, skin, eyes, and kidneys. Chronic UC and CD confer an elevated risk for colorectal carcinoma.^5, 6^ Despite its relatively low mortality rate, the extended duration of IBD treatment imposes substantial burdens on both individuals and society.^7-9^

Initially, IBD was primarily regarded as a condition prevalent in Western regions due to its heightened occurrence there.^10, 11^ In Europe, the diagnosis of IBD affects up to 2 million individuals, and in the United States, there are approximately 1.6 million IBD patients, comprising 785,000 with CD and 910,000 with UC.^12^ Nevertheless, recent epidemiological studies have unveiled a persistent rise in the incidence of IBD within developing nations.^13^ Notably, the newly industrialized countries in Asia and Latin America are currently undergoing an accelerated phase of IBD evolution, potentially edging closer to the incidence rate observed in Western nations within the next decade.^14^

The pathogenesis of IBD is intertwined with factors such as genetic susceptibility of the host, intestinal microbiota, environmental influences, and immune aberrations.^15^ The interaction of genetic susceptibility and environmental factors in the microbiome contributes to the development of IBD by instigating inappropriate immune activation through weakening the intestinal barrier.^16, 17^ The process of industrialization in developing countries has resulted in increasingly Westernized lifestyle, characterized by dietary patterns rich in fats and refined sugars, increased smoking, decreased breastfeeding, and frequent antibiotic exposure.^18, 19^ Individuals with genetic susceptibility mutations are exposed to environmental factors associated with social westernization that may alter their intestinal environment, thereby driving an increase in the incidence of IBD.^20^

Each phase in the evolution of IBD presents distinct clinical challenges.^21^ Currently, most literature concerning the disease burden of IBD primarily focuses on Western countries, leaving developing and newly industrialized countries with limited coverage.^22^ The Asian region encompasses a multitude of developing countries at varying stages of IBD progression. Therefore, comprehending the current epidemiological characteristics of the disease in this region becomes imperative, enabling countries and locales in Asia to proactively address the evolving IBD burden in the upcoming several decades. Based on the latest Global Burden of Diseases, Injuries, and Risk Factors Study (GBD) 2019, we conducted an analysis of the IBD burden in Asia spanning from 1990 to 2019. Furthermore, we project the anticipated incidence trends of IBD in the region for 20 years.

## Methods

### Overview

The GBD 2019 provides a standardized approach to estimate incidence, prevalence, mortality, years of life lost (YLLs), years lived with disability (YLDs), and disability-adjusted life-years (DALYs) across 369 diseases and injuries in 204 countries and territories and for both genders. All data is derived from censuses, household surveys, civil registration and vital statistics, disease registries, use of health services, air pollution monitoring, satellite imaging, disease notification, and other sources. Detailed descriptions of the overall methodology, definitions, and statistical modeling used in GBD 2019 have been described in previous publications.^23, 24^ All data from GBD 2019 comply with the Guidelines for Reporting Health Estimates (GATHER) statement.^25, 26^

We extracted annual metrics encompassing all age groups and age-standardized values for various measures of IBD burden, along with their corresponding 95% uncertainty intervals (UIs), for the period spanning 1990 to 2019 in the Asian region. These data were obtained from the GBD database through the Global Burden of Disease Study 2019 (GBD 2019) Results platform^42^.

The variables obtained from the GBD include the number and percentage of incidence, prevalence, deaths, YLLs, YLDs, DALYs, and their corresponding age-standardized rates (ASR) in the country and Asian region. For clarity, the definitions of the calculated metrics are as follows: the incidence rate (per 100,000) was computed as the number of new cases divided by the population size; the prevalence rate (per 100,000) was calculated by aggregating new cases and previously diagnosed cases, then dividing by the population size; the death rate (per 100,000) was derived from the annual count of deaths divided by the total population size; YLLs were determined by summing the product of each death count, standard life expectancy at each age, and DALYs were computed as the sum of YLLs and YLDs. These extracted metrics from the GBD database serve as valuable resources for assessing and analyzing the evolving burden of IBD across time in the Asian context.

### EAPC

The Estimated Average Percentage Change (EAPC) was employed to evaluate trends in the Age-Standardized Rate (ASR) of incidence over a defined timeframe. This method assumes a consistent and continuous annual variation of the ASR throughout the observed period.^27^ A regression line was fitted to the natural logarithm of the rate i.e., *y = α* + *βx* + *ε, y* = *ln* (*ASR*), x = calendar year, and ε = error term. The EAPC was calculated as 100 × (*exp* (*β*) - 1) and its 95% uncertainty interval (UI) can also be calculated from the linear regression model. When both the EAPC estimation and the lower limit of its 95% UI are greater than zero, it indicates that the ASR is experiencing an upward trend. Conversely, if the EAPC estimation and the upper limit of its 95% UI are less than zero, indicating a declining trend of ASR. In cases where neither of these conditions is met, the ASR is assumed to remain stable over the observed period.^28, 29^ A *p <* 0.05 was considered statistically significant.

### Prediction of IBD burden

To project the number of new cases by location, sex, and age from 2019 to 2040, a log-linear age-period-cohort model is fitted to the recent trends, which can level off the exponential growth and constrain the linear trend prediction. The model is implemented in R via the NORDPRED package.^30^ The extrapolation over the last 3, 4, or 5-year observation periods was performed using a power function to level off the growth and predict the recent linear trend over the last decade. The trend weakens (or strengthens in the case of negative trends) by 25%, 50%, and 75%, respectively, in the second, third, and fourth forecast periods. The number of new cases in 2040 is projected by taking the weighted average of the projected incidence for the last two forecast periods, centered on 2040, and applying these to population forecast data for that year.^31,43^

### Decomposition Analysis of IBD burden

Using decomposition analysis to understand explanatory factors for drove changes in IBD incidence and DALYs between 1990 and 2019. Decomposition analysis is an analytical method to determine the additive contribution of factor differences between two populations to their overall value differences. Decomposing the incidence and DALYs of IBD by age group, population growth, and epidemiological changes allows for the quantification of the individual contributions of these factors to the overall outcome. Additional details are provided in the supplementary methods.^32^

### Frontier Analysis of IBD DALYs

To assess the relationship between the burden of IBD and socio-demographic index (SDI), we applied frontier analysis as a quantitative method to determine the lowest achievable age-standardized DALYs rate based on the developmental status of SDI measurements. SDI is a comprehensive indicator of socio-demographic development, representing the degree of development associated with health outcomes within a particular geographic area. Additional details are provided in the supplementary methods.^32^ All statistics were processed using R Studio (version 4.3.1).

### Patient and public involvement

The present study was a retrospective analysis of chronic disease data, conducted without patient involvement in the design and execution.

## Results

### IBD burden in Asia

In Asia, the total number of IBD incidence cases increased from 64,768 (95% UI 54,186-76,819) in 1990 to 145,561 (95% UI 124,960-170,895) in 2019, an increase of 124.7% **(Table 1)**. Qatar has seen the largest increase, up to 862.7%, followed by the United Arab Emirates with an increase of 793.6%. ASR exhibited an average annual increase of 0.78% (95% UI 0.69%-0.86%) during the same timeframe (from 2.3 per 100,000 in 1990 to 2.9 per 100,000 in 2019) **(Table 1)**. Taiwan (China) exhibited the most substantial increase in ASR at 140.57% (EAPC = 3.87; 95% UI 3.48-4.27), trailed by the Republic of Korea with an EAPC of 1.94 (95% UI 1.59-2.29), and Viet Nam with an EAPC of 2.62 (95% UI 2.13-3.11). Only Georgia, Mongolia, Singapore, and Yemen showed a downward trend in the ASR of IBD in Asia from 1990 to 2019. The incidence cases and the age-standardized incidence rates were higher in males than in females during this period **(Figure 1)**. In 2019, the number of female incidence cases was 62,840 (95% UI 54,255-73,496), while the number of male incidence cases increased to 82,721 (95% UI 70,654-97,174), accounting for 56.83% of the total number of IBD incidence cases. In 2019, the age-standardized incidence rate was 3.31 (95% UI 2.85-3.38) per 100,000 population for males and 2.56 (95% UI 2.21-2.98) per 100,000 population for females.

**Figure 1.**
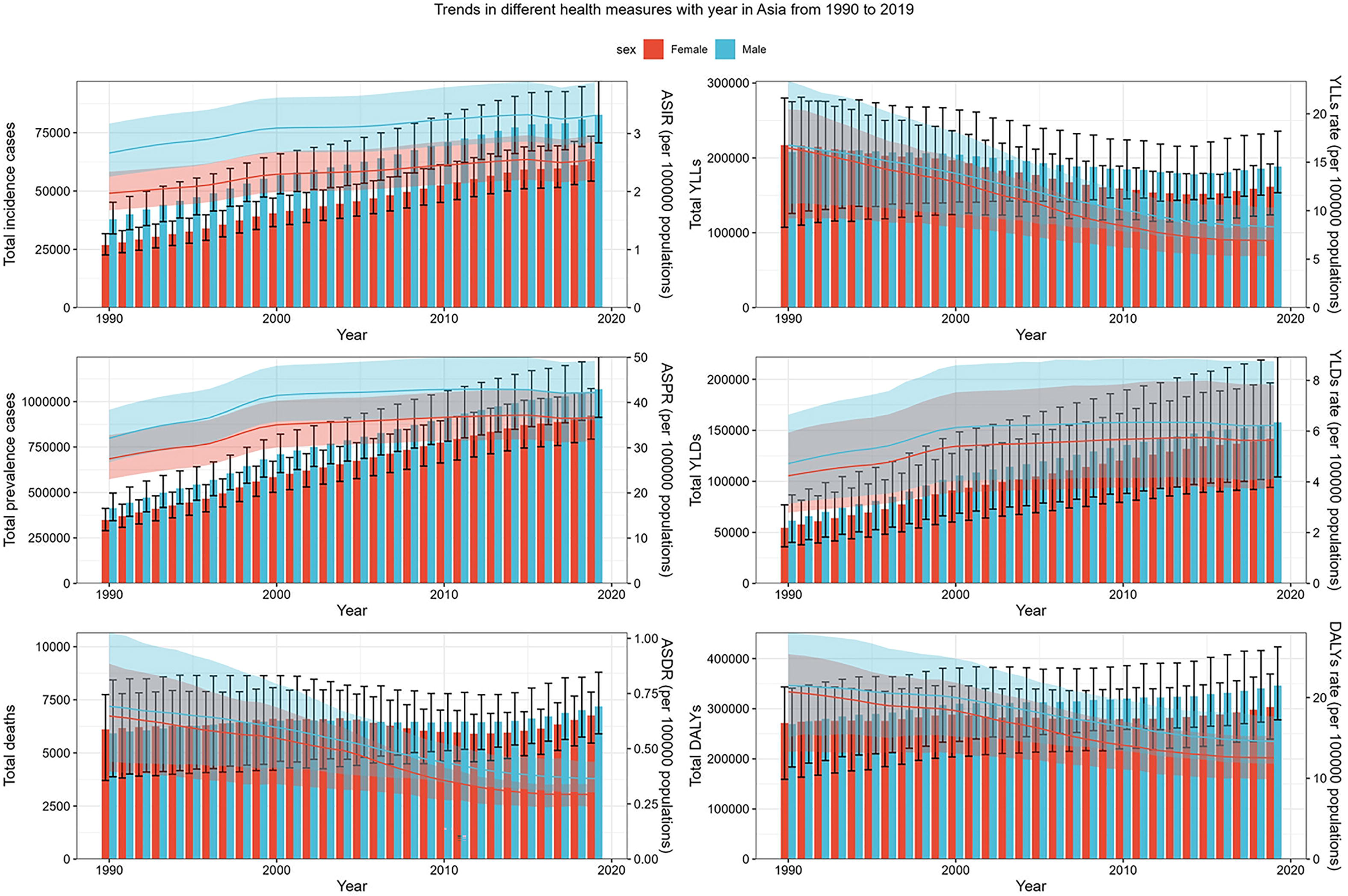
Trends in different health measures of IBD with the year in Asia from 1990 to 2019. Error bars indicate the 95% uncertainty interval (UI) for different measure cases. Shading indicates the 95% UI for the age-standardized rate. IBD, inflammatory bowel disease; ASIR, age-standardized incidence rate; ASPR, age-standardized prevalence rate; ASDR, age-standardized deaths rate. YLLs, years of life lost; YLDs, years lived with disability; DALYs, disability-adjusted life-years.

**Table 1.** The incidence cases, age-standardized incidence, and temporal trends of IBD in 1990 and 2019.

| Location | 1990 |  | 2019 |  | 1990-2019 |
| --- | --- | --- | --- | --- | --- |
|  | Incidence cases | ASR per 100,000 | Incidence cases | ASR per 100,000 | EAPC |
|  | No.×10<sup>2</sup> (95% UI) | No. (95% UI) | No.×10<sup>2</sup> (95% UI) | No. (95% UI) | No. (95% UI) |
| Asia | 647.7 (541.9-768.2) | 2.3 (2.0-2.8) | 1456 (1250-1709) | 2.9 (2.5-3.4) | 0.78 (0.69-0.86) |
| Central Asia | 35.14 (29.95-41.46) | 6.0 (5.1-7.1) | 64.61 (54.87-76.37) | 6.9 (5.9-8.1) | 0.50 (0.46-0.53) |
| Armenia | 1.78 (1.5-2.09) | 5.5 (4.6-6.5) | 2.37 (2.00-2.83) | 6.6 (5.6-8.0) | 0.71 (0.66-0.76) |
| Azerbaijan | 4.32 (3.65-5.10) | 6.7 (5.7-8.0) | 8.05 (6.77-9.65) | 7.0 (5.9-8.4) | 0.18 (0.13-0.23) |
| Georgia | 3.81 (3.25-4.49) | 6.5 (5.5-7.6) | 2.75 (2.34-3.27) | 6.3 (5.4-7.4) | -0.16 (-0.21-0.12) |
| Kazakhstan | 9.22 (7.82-10.89) | 6.0 (5.1-7.1) | 13.89 (11.85-16.6) | 7.2 (6.1-8.6) | 0.43 (0.31-0.56) |
| Kyrgyzstan | 2.22 (1.89-2.60) | 6.0 (5.1-7.1) | 3.90 (3.29-4.60) | 6.4 (5.4-7.5) | 0.12 (0.06-0.18) |
| Mongolia | 0.98 (0.82-1.16) | 6.2 (5.3-7.5) | 2.51 (2.10-3.00) | 7.4 (6.2-8.7) | 0.58 (0.55-0.61) |
| Tajikistan | 2.25 (1.90-2.67) | 5.8 (4.9-6.9) | 5.18 (4.39-6.12) | 6.2 (5.3-7.3) | 0.16 (0.10-0.22) |
| Turkmenistan | 1.88 (1.57-2.22) | 6.8 (5.7-8.0) | 3.83 (3.24-4.51) | 7.7 (6.5-9.0) | 0.54 (0.44-0.63) |
| Uzbekistan | 8.68 (7.32-10.30) | 5.4 (4.6-6.5) | 22.13 (18.78-26.42) | 6.9 (5.9-8.2) | 1.04 (0.96-1.13) |
| East Asia | 175.67 (146.3-208.02) | 1.4 (1.2-1.7) | 523.19 (446.8-614.55) | 3.0 (2.5-3.4) | 2.54 (2.40-2.68) |
| China | 172.2 (143.3-204) | 1.5 (1.2-1.7) | 514.6 (439.3-604.7) | 3.0 (2.6-3.5) | 2.54 (2.40-2.68) |
| Japan | 163.8 (138.3-195.7) | 11.2 (9.5-13.3) | 224.4 (193.5-259.1) | 19.6 (16.9-22.7) | 1.32 (1.03-1.60) |
| North Korea (DPRK) | 1.97 (1.64-2.36) | 1.0 (0.8-1.1) | 3.87 (3.26-4.63) | 1.3 (1.1-1.5) | 1.23 (1.08-1.38) |
| Republic of Korea | 15.87 (13.9-17.77) | 3.4 (3.0-3.8) | 44.07 (42.2-45.89) | 7.3 (7.0-7.6) | 1.94 (1.59-2.29) |
| Taiwan (China) | 1.49 (1.29-1.71) | 0.7 (0.6-0.8) | 4.70 (4.2-5.26) | 1.7 (1.5-1.9) | 3.87 (3.48-4.27) |
| Middle East and North Africa | 81.07 (70.4-92.04) | 2.9 (2.6-3.4) | 227.2 (192.77-269.52) | 3.7 (3.2-4.5) | 0.62 (0.53-0.72) |
| Afghanistan | 1.92 (1.59-2.28) | 2.3 (1.9-2.7) | 6.84 (5.57-8.28) | 2.4 (2.0-3.0) | 0.28 (0.17-0.39) |
| Bahrain | 0.13 (0.11-0.15) | 2.6 (2.3-3.0) | 0.58 (0.48-0.72) | 3.2 (2.7-3.9) | 0.45 (0.29-0.61) |
| Iran (Islamic Republic of) | 13.47 (11.11-16.54) | 3.3 (2.7-4.1) | 31.67 (25.99-38.94) | 3.4 (2.9-4.2) | 0.21 (0.06-0.35) |
| Iraq | 2.40 (2.02-2.82) | 1.8 (1.5-2.1) | 9.84 (8.10-11.91) | 2.5 (2.1-3.1) | 1.33 (1.19-1.46) |
| Jordan | 1.30 (1.14-1.47) | 4.4 (3.9-5.0) | 7.79 (6.49-9.17) | 6.9 (5.8-8.1) | 1.49 (1.34-1.65) |
| Kuwait | 0.61 (0.57-0.66) | 3.4 (3.2-3.7) | 1.95 (1.61-2.39) | 3.7 (3.1-4.5) | 0.41 (0.30-0.51) |
| Lebanon | 0.65 (0.54-0.80) | 2.3 (1.9-2.9) | 1.94 (1.61-2.38) | 3.6 (3.0-4.4) | 1.39 (1.29-1.49) |
| Oman | 0.38 (0.31-0.46) | 2.4 (2.0-2.9) | 1.43 (1.15-1.73) | 2.7 (2.3-3.3) | 0.06 (-0.11-0.22) |
| Palestine | 0.29 (0.24-0.35) | 2.0 (1.7-2.4) | 1.22 (1.01-1.45) | 2.9 (2.5-3.5) | 1.11 (0.93-1.29) |
| Qatar | 0.13 (0.10-0.16) | 2.8 (2.4-3.4) | 1.25 (1.02-1.52) | 3.6 (3.0-4.3) | 0.90 (0.83-0.96) |
| Saudi Arabia | 3.41 (2.87-4.04) | 2.6 (2.2-3.1) | 11.22 (9.29-13.53) | 2.7 (2.2-3.2) | -0.14 (-0.28-0) |
| Syrian Arab Republic | 2.41 (2.00-2.89) | 2.7 (2.2-3.3) | 4.49 (3.76-5.44) | 3.2 (2.6-3.8) | 0.25 (0.06-0.45) |
| Turkey | 20.84 (18.59-23.16) | 3.9 (3.5-4.3) | 59.61 (51.19-68.93) | 6.4 (5.5-7.4) | 1.27 (1.07-1.47) |
| United Arab Emirates | 0.58 (0.47-0.71) | 3.2 (2.7-3.9) | 5.20 (4.10-6.51) | 4.1 (3.4-5.0) | 0.76 (0.60-0.93) |
| Yemen | 1.92 (1.57-2.30) | 2.1 (1.7-2.6) | 6.47 (5.27-7.93) | 2.5 (2.1-3.1) | 0.73 (0.69-0.77) |
| South Asia | 184.39 (151.69-225.8) | 2.2 (1.8-2.6) | 397.1 (329.43-484.54) | 2.3 (1.9-2.8) | 0.47 (0.32-0.62) |
| Cambodia | 0.30 (0.23-0.37) | 0.4 (0.3-0.5) | 0.79 (0.64-0.96) | 0.5 (0.4-0.6) | 1.06 (1.01-1.11) |
| Bangladesh | 13.55 (11.05-16.56) | 1.8 (1.5-2.2) | 34.04 (27.88-41.36) | 2.2 (1.8-2.6) | 0.70 (0.62-0.77) |
| Bhutan | 0.07 (0.06-0.09) | 1.7 (1.4-2.1) | 0.14 (0.12-0.17) | 1.9 (1.6-2.3) | 0.09 (-0.1-0.28) |
| India | 153.6 (126.31-188.15) | 2.2 (1.9-2.7) | 317.8 (264.33-389.72) | 2.3 (1.9-2.9) | 0.42 (0.24-0.61) |
| Nepal | 2.42 (1.98-3.01) | 1.7 (1.4-2.1) | 6.24 (5.15-7.64) | 2.2 (1.8-2.8) | 1.03 (0.96-1.09) |
| Pakistan | 14.74 (12.18-17.92) | 1.9 (1.6-2.3) | 38.88 (31.93-48.38) | 2.3 (1.9-2.9) | 0.69 (0.65-0.74) |
| Southeast Asia | 18.82 (15.27-22.7) | 0.5 (0.4-0.6) | 49.8 (41.82-59.72) | 0.7 (0.6-0.8) | 1.45 (1.28-1.62) |
| Brunei Darussalam | 0.13 (0.10-0.15) | 5.0 (4.1-6.0) | 0.26 (0.21-0.31) | 5.1 (4.2-6.1) | 0.08 (0.04-0.12) |
| Indonesia | 6.81 (5.51-8.28) | 0.4 (0.4-0.5) | 15.04 (12.48-18.08) | 0.6 (0.5-0.7) | 0.85 (0.77-0.93) |
| Lao PDR | 0.12 (0.10-0.15) | 0.4 (0.3-0.5) | 0.35 (0.29-0.43) | 0.5 (0.4-0.6) | 1.04 (0.98-1.09) |
| Malaysia | 0.72 (0.61-0.83) | 0.5 (0.4-0.5) | 2.51 (2.23-2.83) | 0.8 (0.7-0.9) | 1.60 (1.48-1.73) |
| Maldives | 0.01 (0-0.01) | 0.3 (0.3-0.4) | 0.03 (0.02-0.03) | 0.5 (0.4-0.6) | 1.26 (1.18-1.34) |
| Myanmar | 1.41 (1.13-1.73) | 0.4 (0.3-0.5) | 3.06 (2.53-3.78) | 0.6 (0.5-0.7) | 1.22 (1.15-1.29) |
| Philippines | 2.42 (1.97-2.92) | 0.5 (0.4-0.6) | 6.57 (5.41-7.99) | 0.6 (0.5-0.8) | 0.78 (0.69-0.88) |
| Singapore | 1.23 (1.01-1.47) | 3.5 (2.9-4.1) | 2.15 (1.78-2.64) | 3.2 (2.7-3.9) | -0.35 (-0.56--0.14) |
| Sri Lanka | 1.07 (0.88-1.31) | 0.7 (0.6-0.8) | 2.68 (2.39-3.01) | 1.1 (1.0-1.3) | 1.86 (1.63-2.10) |
| Thailand | 1.83 (1.42-2.30) | 0.3 (0.3-0.4) | 3.73 (3.09-4.55) | 0.5 (0.4-0.6) | 1.11 (1.03-1.18) |
| Timor-Leste | 0.02 (0.02-0.03) | 0.4 (0.3-0.4) | 0.06 (0.05-0.07) | 0.5 (0.4-0.6) | 1.50 (1.36-1.64) |
| Viet Nam | 4.02 (3.16-4.99) | 0.7 (0.5-0.8) | 14.82 (12.01-18.36) | 1.4 (1.1-1.7) | 2.62 (2.13-3.11) |

ASR, age-standardized rate (per 100 000 population); EAPC, estimated annual percentage change; UI, uncertainty interval. DPRK, Democratic People's Republic of Korea; Lao PDR, Lao People's Democratic Republic.

Between 1990 and 2019, the number of patients with IBD in Asia increased from 0.76 million (95% UI 0.63-0.91) to over 1.99 million (95% UI 1.71-2.32), an increase of 160.92% **(Supplementary Table 1)**. Meanwhile, the age-standardized prevalence rate increased from 29.8 (95% UI 24.9-35.5) per 100,000 population in 1990 to 39.4 (95% UI 33.7-45.8) per 100,000 population in 2019 **(Supplementary Table 2)**. The number of prevalence cases also showed a gender difference, with the total number of prevalence cases of IBD increasing from 414,592 (95% UI 344,796-497,045) to 1,068,534 (95% UI 913,134-1,246,870) in males and from 348,952 (95% UI 289,914-412,408) to 923,687 (95% UI 793,214-1,072,032) in females. The total number of prevalence cases and the age-standardized prevalence rate of IBD in Asia in 2019 are shown in **Figure 2**. China (911,045, 95% UI 776,347-1,069,533) has the highest number of IBD prevalence cases in Asia, followed by Japan (408,632, 95% UI 468,152-353,858) and India (270,719, 95% UI 332,264-219,873). While the countries with the highest age-standardized prevalence rate were Japan (291.9, 95% UI 251.8-336.6) and Jordan (113.5, 95% UI 95.8-133.9), the lowest were Lao People’s Democratic Republic (3.7, 95% UI 3.0-4.6) and Myanmar (3.7, 95% UI 3.0-4.6) **(Supplementary Table 3)**.

**Figure 2.**
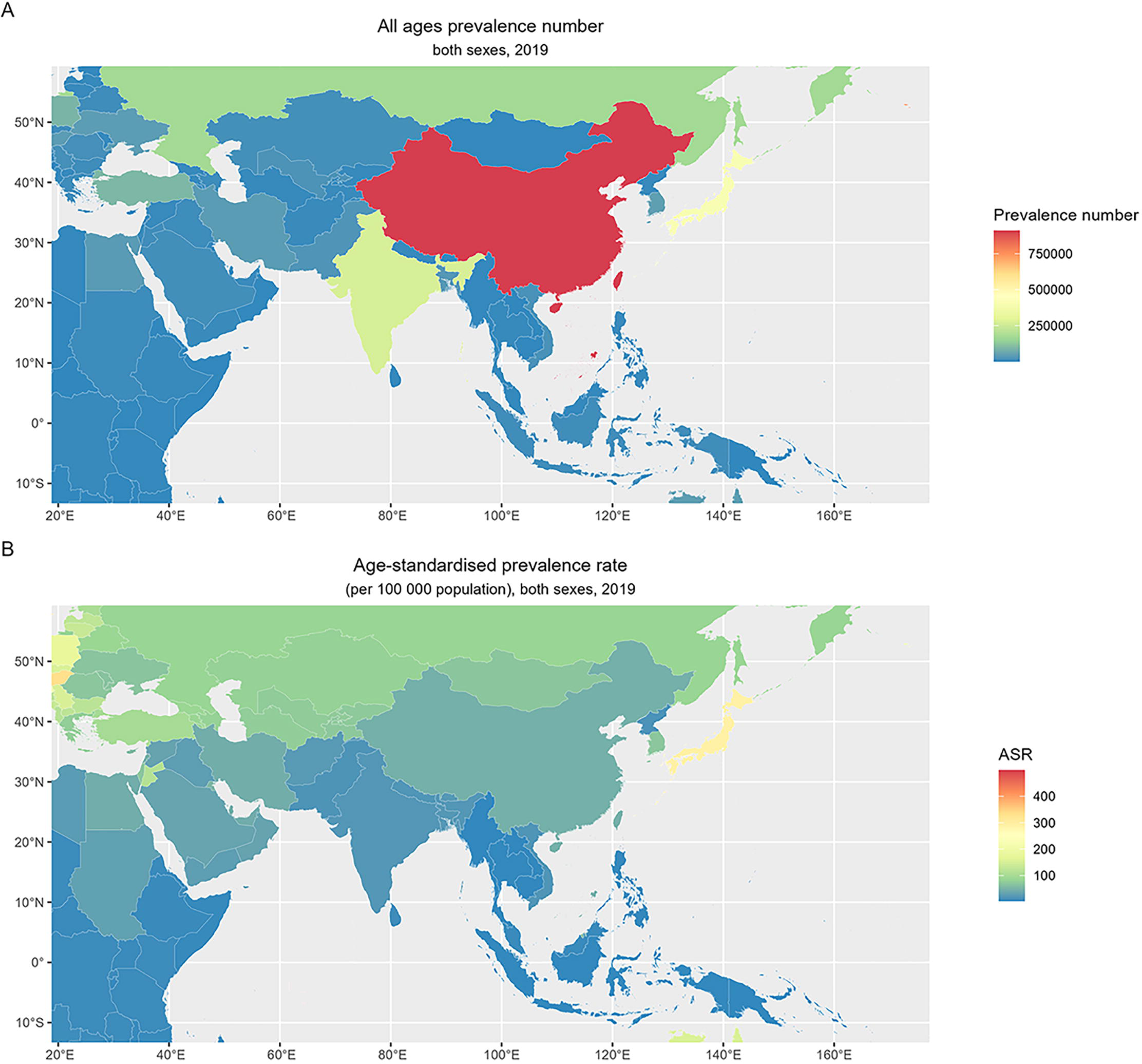
The total number and age-standardized rates of prevalence of IBD in Asia. (A) The total number of prevalence cases of IBD in Asia, both sexes, for 50 countries and territories, 2019. (B) The age-standardized prevalence rate of IBD, both sexes, for 50 countries and territories, 2019. IBD, inflammatory bowel disease; ASR, age-standardized rate.

From 1990 to 2019, the total number of IBD-related deaths in Asia increased by 16.03%, from 12,030 (95% UI 8,669-15,526) to 13,957 (95% UI 11,898-16,021) **(Figure 1)**. Although the increase in the total number of deaths, the age-standardized death rate decreased by 51.50%, from 0.67 (95% UI 0.46-0.90) per 100,000 population to 0.33 (95% UI 0.28-0.37) per 100,000 population. There was no significant difference in gender among the total IBD-related deaths, with females initially experiencing more deaths than males, but males gradually overtaking females after 2006. In 2019, China (4676, 95% UI 3774-5461) had the highest total number of deaths due to IBD, while Brunei Darussalam (1.48, 95% UI 0.97-1.89) had the highest age-standardized death rate in Asia **(Supplementary Table 3)**.

In 1990, the total number of YLLs attributed to IBD was 424,966 (95% UI 278,705-534,164), which was reduced to 350,097 (95% UI 292,031-409,529) in 2019 **(Figure 1)**. Age-standardized YLLs rate for both sexes has decreased over time, with the age-standardized YLLs rate for males decreasing from 16.8 (95% UI 10.7-23.4) in 1990 to 8.3 (95% UI 6.8-10.3) per 100,000 population in 2019, and the age-standardized YLLs rate for females decreasing from 16.5 (95% UI 9.2-20.4) to 6.9 (95% UI 5.3-8.2) per 100,000 population. In 2019, India had the highest total YLLs of any country or region with 122,827 (95% UI 81,271-163,422); the lowest total YLLs were found in Seychelles (95% UI 5.8, 4.3-7.6). Brunei Darussalam had the highest age-standardized rate of YLLs among all countries or regions at 24.1 (95% UI 19.5-29.9) per 100,000 population **(Supplementary Table 4)**.

The total number of YLDs caused by IBD doubled during this period, increasing from 115,982 (95% UI 75,838-162,919) in 1990 to 299,663 (95% UI 198,365-418,635) in 2019 **(Figure 1)**. In addition, the age-standardized YLDs rate increased from 4.5 (95% UI 3.0-6.3) per 100,000 population in 1990 to 5.9 (95% UI 3.9-8.3) per 100,000 population in 2019. The overall level of total YLDs and the age-standardized YLDs rate caused by IBD was higher in males than in females between 1990 and 2019. The countries with the highest total YLDs caused by IBD in 2019 were China (135,906, 95% UI 89,067-191,529) and India (60,744, 95% UI 40,135-83,792). Japan and Jordan had the highest age-standardized YLDs rate at 44.1 (95% UI 29.1-61.2) and 16.8 (95% UI 11.1-24.2) per 100,000 population in 2019, respectively **(Supplementary Table 4)**.

The total DALYs have increased from 540,949 (95% UI 382,320-666,324) in 1990 to 649,760 (95% UI 530,395-783,181) in 2019, but age-standardized DALYs rate decreased from 21.1 (95% UI 15.8-26.0) per 100,000 population to 13.5 (95% UI 11.1-16.2) per 100,000 population **(Figure 1)**. Total YLLs and YLDs accounted for 53.9% and 46.1% of the total DALYs in 2019, respectively. There is no doubt that China, India, and Japan are the countries with the most severe burden of IBD in Asia, with the total DALYs in 2019 being 232,464 (95% UI 179,903-291,090), 164,416 (95% UI 117,527-208,276) and 67,424 (95% UI 46,877-91,495) respectively. The United Arab Emirates experienced the largest increase in the total DALYs, from 144 (95% UI 93-219) in 1990 to 1401 (95% UI 971-1965) in 2019, which has been an increase of 871.11%. The country with the highest age-standardized DALYs rate in 2019 remains Japan (95% UI 46.6, 31.4-63.5), while the lowest is Thailand (95% UI 3.6, 2.8-4.7) **(Supplementary Table 4)**.

### SDI region

For high SDI regions, the countries with the highest age-standardized incidence rates (per 100,000 population) were Japan (19.6, 95% UI 16.9-22.7) and the Republic of Korea (7.3, 95% UI 7.0-7.6), and the lowest were Seychelles (0.6, 95% UI 0.5-0.7) and Mauritius (0.6, 95% UI 0.5-0.7). It is worth noting that the country with the highest age-standardized death rate (per 100,000 population) is Brunei Darussalam (1.5, 95% UI 1.0-1.9), while Japan characterized by a high age-standardized incidence rate has an extremely low age-standardized death rate (0.11, 95% UI 0.09-0.16). Among the medium SDI regions, Turkmenistan and Mongolia had the highest age-standardized prevalence rates, while the Maldives and Thailand had low age-standardized prevalence rates. The highest age-standardized death rates were observed in Indonesia (0.5, 95% UI 0.3-0.7) and Mongolia (0.5, 95% UI 0.3-0.6). In the low SDI region, Tajikistan had the highest age-standardized incidence rate and Cambodia the lowest. Pakistan and Nepal have a higher age-standardized death rates of 0.6 (95% UI 0.4-1.0) per 100,000 population **(Figure 3)**.

**Figure 3.**
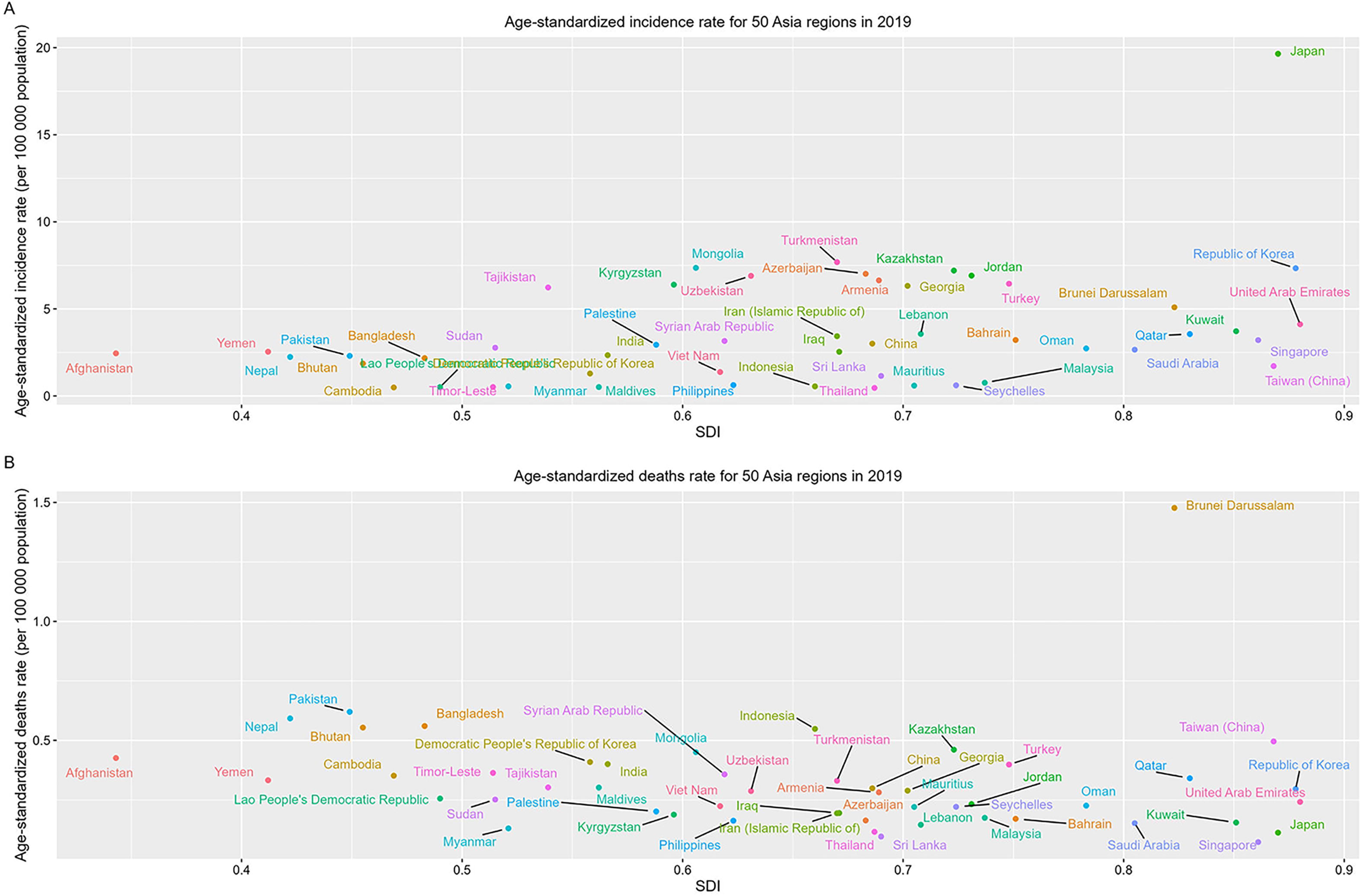
The age-standardized incidence and death rate of IBD for 50 Asia regions by SDI in 2019. (A) The distribution of age-standardized incidence rate of IBD, both sexes, for 50 countries and territories, 2019. (B) The distribution of age-standardized death rate of IBD, both sexes, for 50 countries and territories, 2019. IBD, inflammatory bowel disease; SDI, socio-demographic index.

### Age distribution of IBD

The age of onset of IBD is relatively broad, with patients in almost all age groups. The 2019 data showed that the 30-34 years age group had the largest number of total incidence cases with 17,802 (95% UI 13,376-22,463). In terms of incidence rate, the 40-44 years age group was the highest of all age groups, reaching 5.2 (95% UI 4.0-6.7) per 100,000 population, and the incidence rate of IBD in males exceeded that of females in all age groups. The total number of prevalence cases of IBD peaked in the age group 45-49 years (261,825, 95% UI 215,036-316,099) and then gradually decreased in the older age group. The prevalence rate increased to a maximum of 91.0 (95% UI 76.2-110.4) per 100,000 population in the age group 55-59 years. In 2019, the 80-84 years age group had the highest total number of IBD-related deaths (1,932, 95% UI 1,612-2,254), and more females (991, 95% UI 745-1180) than males (941, 95% UI 764-1194) died for the first time in this age group. The total YLLs were highest in the 65-69 years age group (30,665, 95% UI 25,256-36,643), where the number of males was 17,424 (95% UI 13,186-22,394). The total YLLs of females reached the highest value in the age group 70-74 years (13274, 95% UI 10,149-15,771). The total YLDs was greatest in the 45-49 years age group (39,140, 95% UI 25,859-56,251) and then decreased in all age groups. As with the YLDs, the total DALYs also peaked in the 45-49 years age group (61,662, 95% UI 47,029-79,870), and then gradually declined **(Figure 4)**.

**Figure 4.**
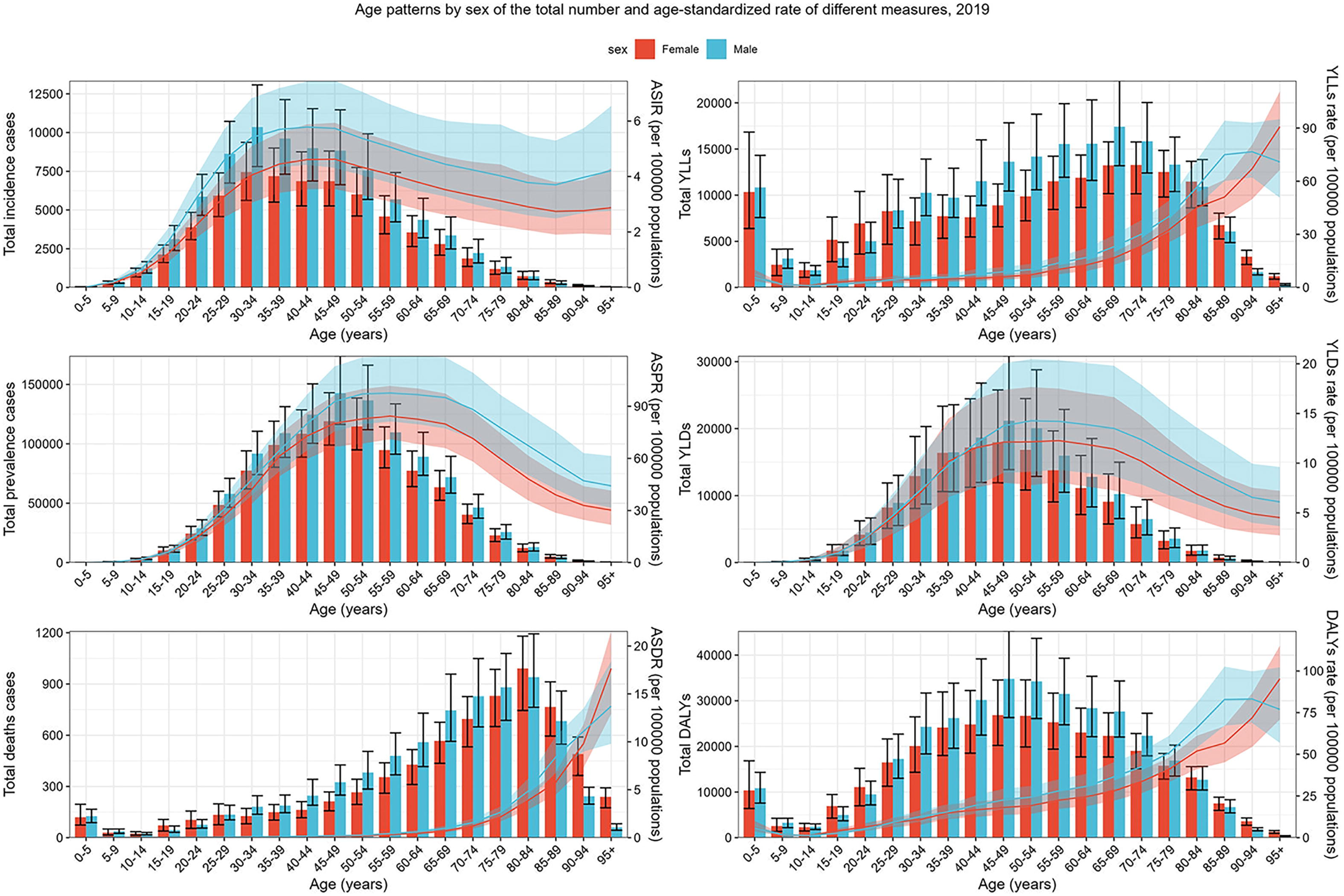
Age patterns by sex in 2019 of the total number and age-standardized rate of different measures of IBD in Asia. Error bars indicate the 95% uncertainty interval (UI) for different measure cases. Shading indicates the 95% UI for the age-standardized rate. IBD, inflammatory bowel disease; ASIR, age-standardized incidence rate; ASPR, age-standardized prevalence rate; ASDR, age-standardized deaths rate. YLLs, years of life lost; YLDs, years lived with disability; DALYs, disability-adjusted life-years.

### Decomposition analysis

To assess the differential impact of aging, epidemiological changes, and population growth on the epidemiology of IBD over a period of approximately 30 years, we performed a decomposition analysis of IBD-related incidence and DALYs in different regions of Asia from 1990 to 2019. In terms of incidence, the number of incidence cases has increased at different rates in Asia and different regions, with the most pronounced increase observed in East Asia and South Asia, and the largest increase in the total number of incidence cases in East Asia over the past 30 years. In Asia, population growth and epidemiologic changes accounted for 43.94% and 30.51% of the increase in IBD incidence cases, respectively. The contribution of population growth to the total incidence was highest in South Asia and the Middle East (68.49% and 63.1%, respectively). The total contribution of population growth was lowest in East Asia, at 17.41%. The contribution of epidemiologic changes to the total number of incidence cases was most evident in East Asia (64.71%), followed by Southeast Asia (43.47%). The increase in the number of IBD-related incidence cases is mainly due to population growth in the South Asia region **(Figure 5-A)**.

**Figure 5.**
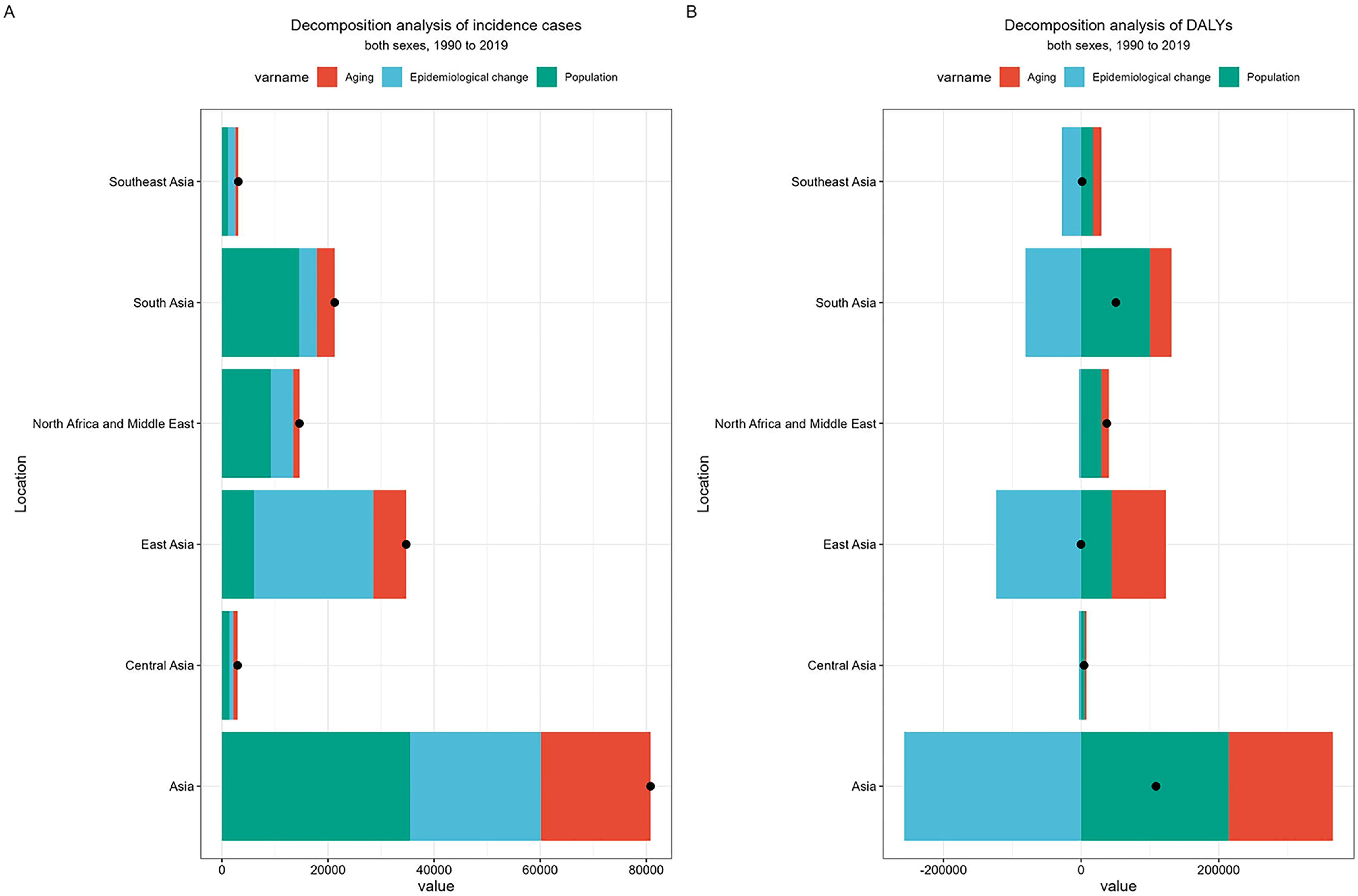
Decomposition analysis of explanatory factors for drove changes in IBD incidence and DALYs in Asia. (A) Changes in the incidence of IBD according to the determinants of population growth, aging, and epidemiological change from 1990 to 2019 at the Asian level. (B) Changes in DALYs of IBD according to the determinants of population growth, aging, and epidemiological change from 1990 to 2019 at the Asian level. The black dots represent the overall change value contributed by the 3 components. The magnitude of a positive value for each component indicates a corresponding increase in the incidence and DALYs of IBD allocated to the component; a negative value of the magnitude indicates a corresponding decrease in the incidence and DALYs of IBD assigned to the related component. IBD, inflammatory bowel disease; DALYs, disability-adjusted life-years.

In terms of DALYs, the overall burden is increasing significantly in Asia. Population growth and aging contributed 197.09% and 139.04%, respectively, to the increase in the burden of DALYs from IBD in Asia, with population growth in South Asia having the greatest impact on the total burden of DALYs. The contribution of aging to total DALYs is most pronounced in East Asia, followed by South Asia. The increase in total DALYs in Asia is driven by the population growth in South Asia and the aging of the population in East Asia, and the overall changes in Central Asia have a smaller impact on the total burden of DALYs **(Figure 5-B)**. In general, population growth has been the major driving force behind the increase in IBD incidence and DALYs burden in the Asian region over the past 30 years. However, several influencing factors do not contribute equally to the increase in the burden of IBD in different regions of Asia, suggesting that the development of IBD may be at different epidemiological stages in different regions of Asia **(Supplementary Table 5)**.

### Frontier analysis

To comprehensively investigate the relationship between the level of development of different countries or regions in Asia and the potential improvement of IBD DALYs, we conducted a frontier analysis of age-standardized DALYs rate and SDI based on IBD development data in Asia from 1990 to 2019 **(Table 2)**. The frontier line depicts the countries and regions with the lowest DALYs rate under SDI conditions, which represents the level of DALYs that should be achieved under that SDI condition. The effective difference refers to the distance between the observed DALYs in a country or region and the achievable DALYs, which are measured as the distance from the dot to the line. The socio-demographic resources of different countries or regions have the potential to reduce or eliminate this gap in the future. Concurrently, the trend of age-standardized DALYs rate in different countries or regions is visually represented by color. If the age-standardized DALYs rate in 2019 is higher than in 1990, it is labeled with “Increase” and shown in blue; conversely, if it is marked as a “Decrease”, it is shown in red **(Figure 6)**.

**Figure 6.**
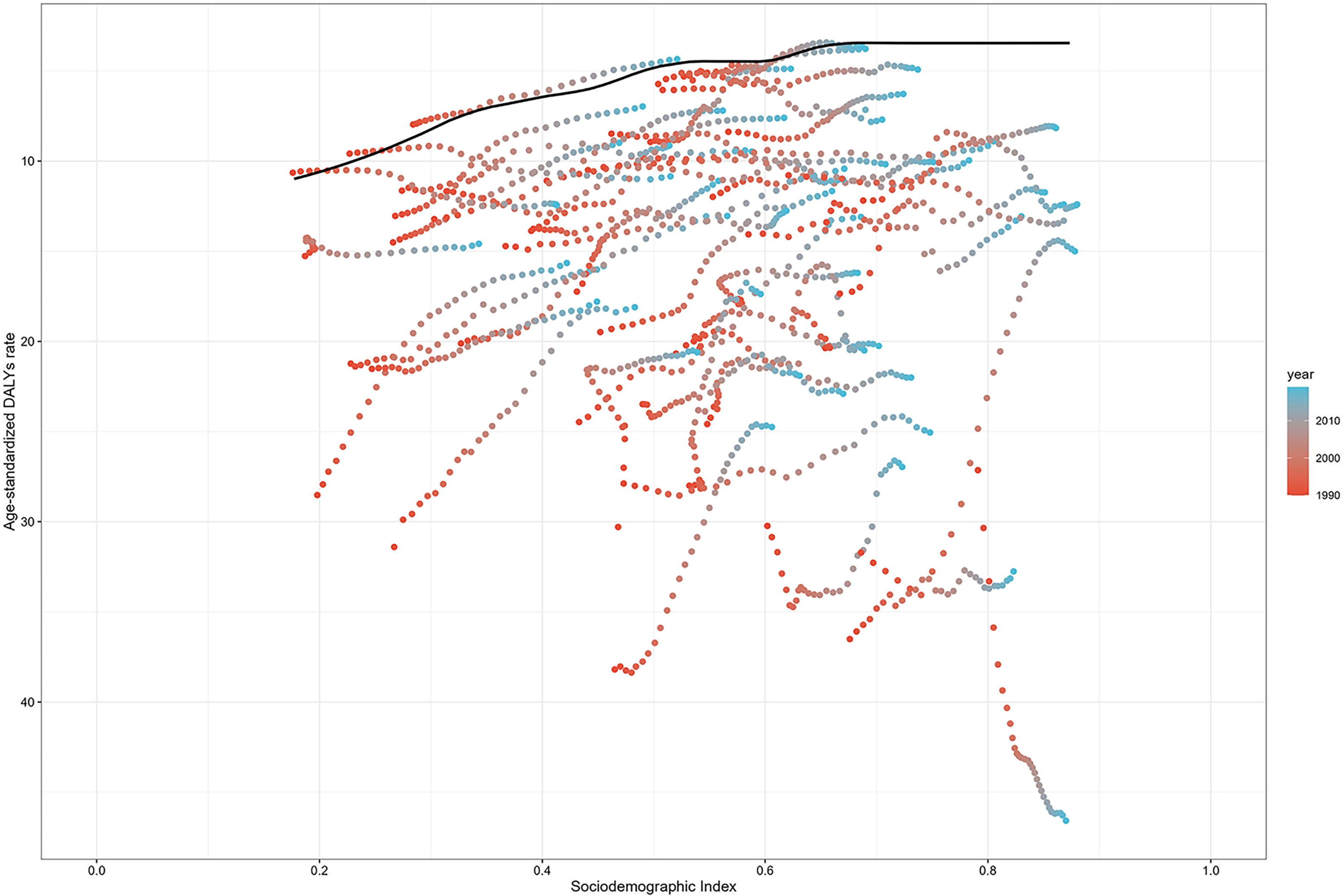
Frontier analysis based on SDI and age-standardized DALYs rate of IBD from 1990 to 2019 in Asia. The color scale represents the years from 1990 depicted in red to 2019 depicted in blue. The frontier is delineated in a solid black color. IBD, inflammatory bowel disease; SDI, socio-demographic index; DALYs, disability-adjusted life-years.

**Table 2.**
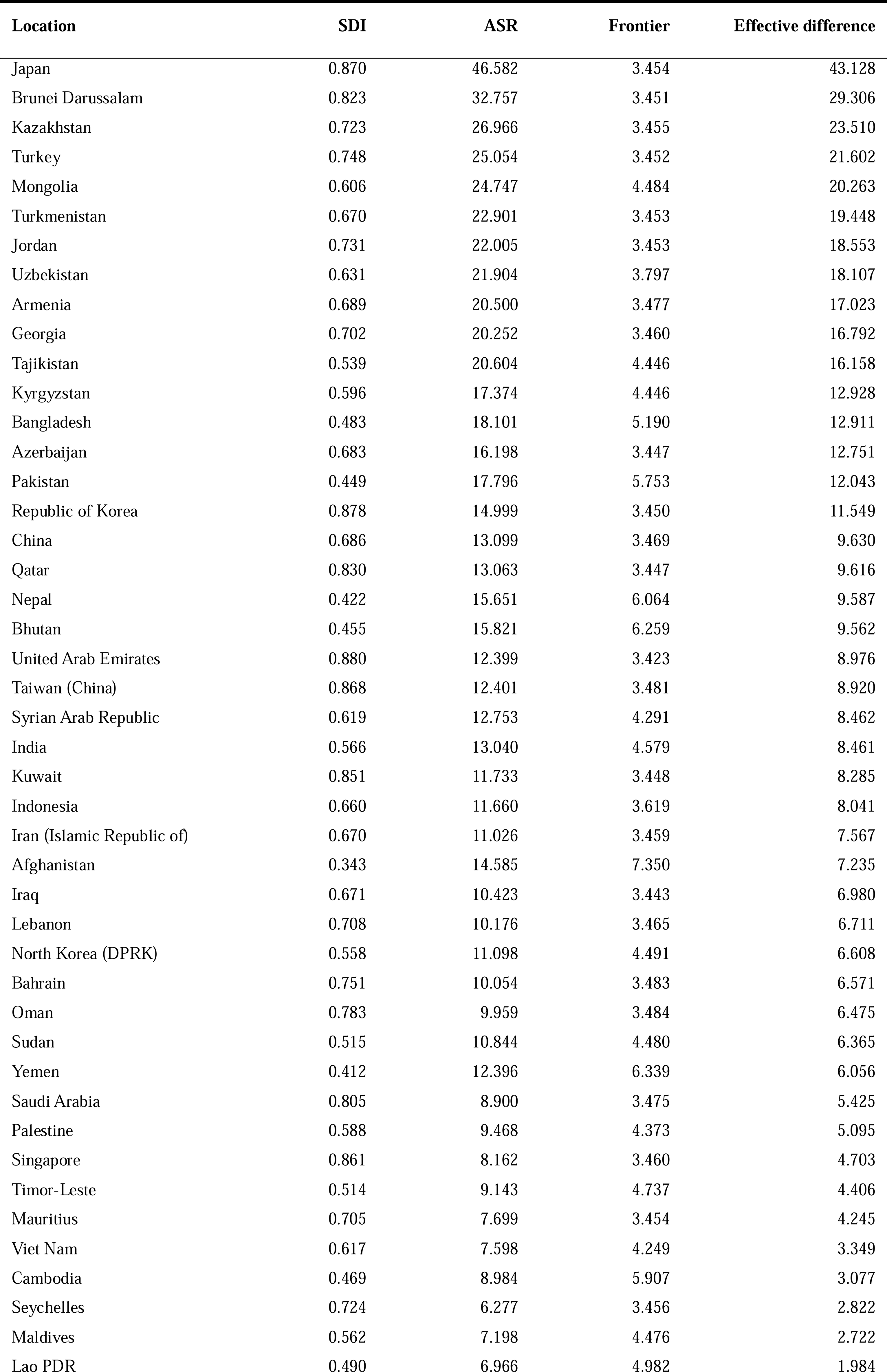
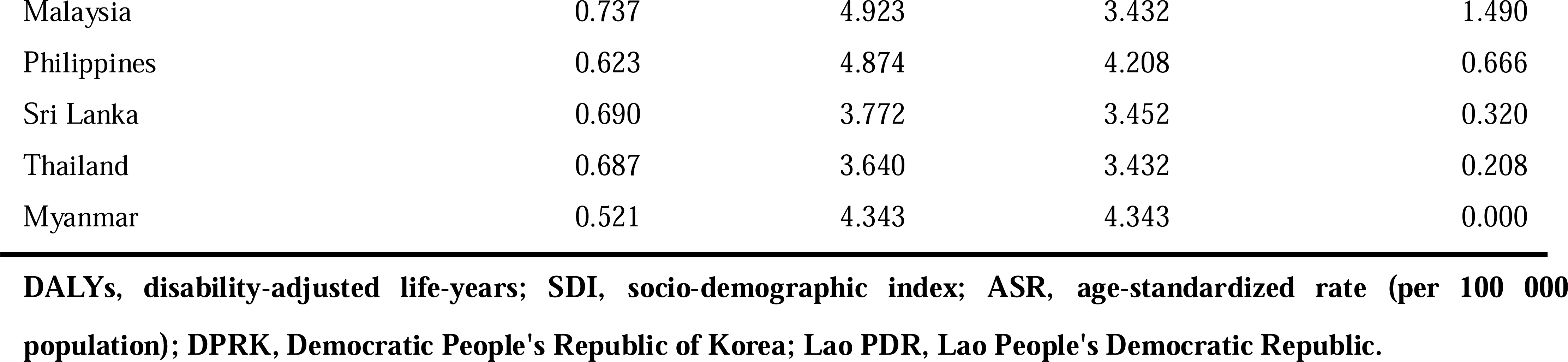
A frontier analysis of age-standardized DALYs rate and SDI based on IBD development data in Asia from 1990 to 2019.

The effective differences were calculated using DALYs and SDI for different countries or regions in the Asian region in 2019. The 14 countries or regions with the largest effective differences from the frontier line are Japan, Brunei Darussalam, Kazakhstan, Turkey, Mongolia, Turkmenistan, Jordan, Uzbekistan, Armenia, Georgia, Tajikistan, Kyrgyzstan, Bangladesh, and Azerbaijan, with effective differences ranging from 12.75 to 43.13; in particular, these countries have significantly higher IBD DALYs than other countries with the same level of SDI. The 5 countries or regions with the smallest effective differences among the low SDI countries or regions are the Lao People’s Democratic Republic, Cambodia, Yemen, Afghanistan, and Bhutan. These countries have relatively low levels of DALYs rate and are the best performers at comparable SDI levels, but Yemen is the only one of these countries where the age-standardized DALYs rate is higher than in 1990. The 5 countries or regions with the largest effective differences in the high SDI are Japan, the Republic of Korea, the United Arab Emirates, Taiwan (China), and Kuwait. They are the worst performing countries or regions at the same SDI level, with the age-standardized DALYs rate of Japan and the United Arab Emirates showing an upward trend compared with 1990 **(Figure 7)**.

**Figure 7.**
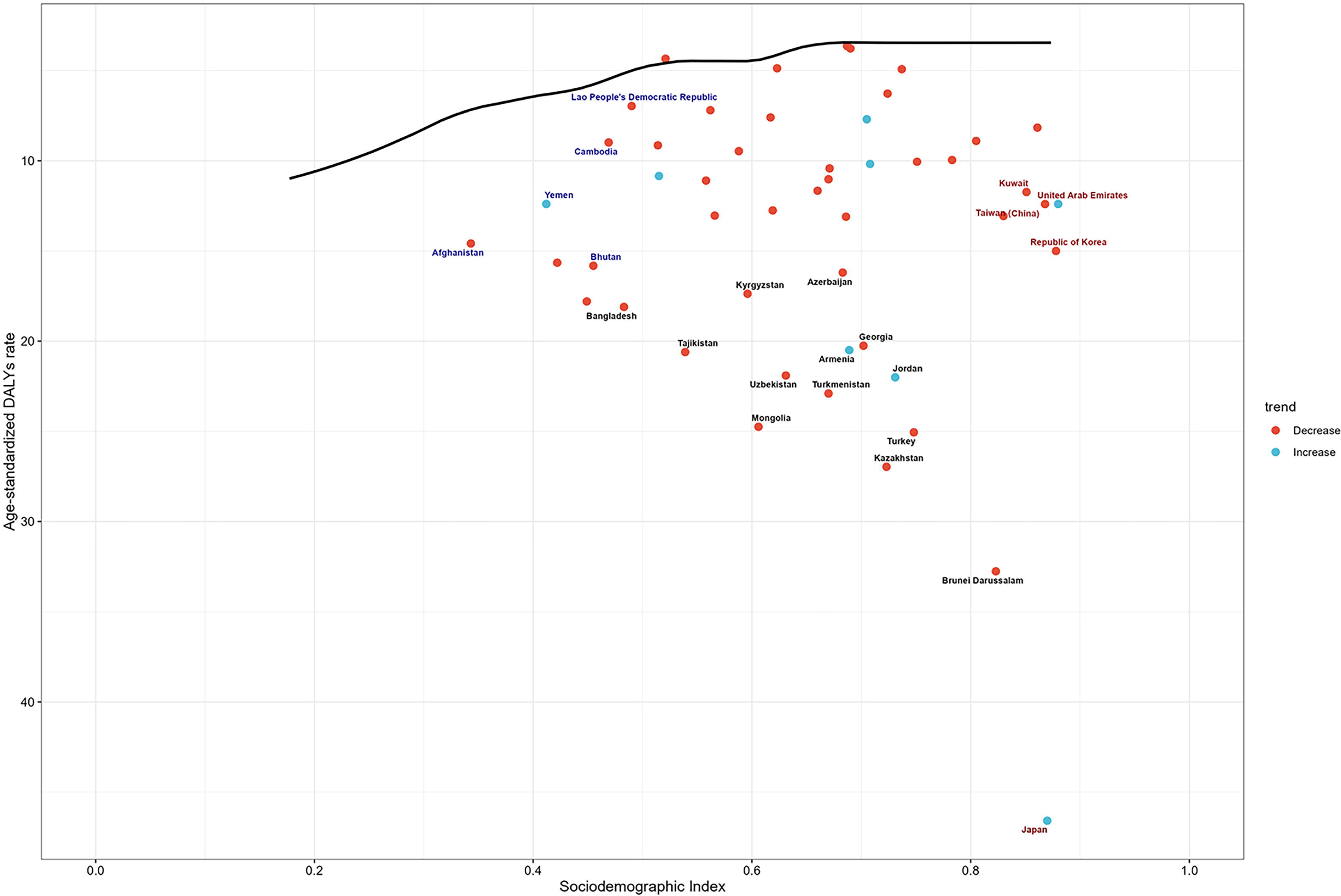
Frontier analysis based on SDI and age-standardized DALYs rate of IBD in 2019. The frontier is delineated in solid black color; countries and territories are represented as dots. The top 14 countries with the largest effective difference are labeled in black; the 5 frontier countries with low SDI (<0.5) and low effective difference are labeled in blue and the 5 countries and territories with high SDI (>0.85) and relatively high effective difference are labeled in red. Blue dots indicate an increase in the age-standardized DALYs rate from 1990 to 2019; red dots indicate a decrease in the age-standardized DALYs rate from 1990 to 2019. SDI, socio-demographic index; DALYs, disability-adjusted life-years; IBD, inflammatory bowel disease.

### Prediction of IBD burden

A review of the trends from 1990 to 2019 shows that the IBD epidemic in Asia is still in a phase of accelerating the incidence rate. Based on previous data, we projected changes in the incidence of IBD in Asia over the next 20 years using the APC model. It is expected that the overall trend of IBD incidence in Asia will continue to increase over the next 20 years, and the number of IBD cases is expected to reach 179,756 in 2040, of which 100,907 cases in males and 788,49 cases in females **(Supplementary Table 6)**. There are similar development trends in the number of incidence cases in males and females, but the total number of cases in males will continue to be higher than in females. The age-standardized incidence rate of IBD continues to increase from 2019 to 2029, and is expected to reach 2.96 per 100,000 population in 2029. The age-standardized incidence rate then shows a downward trend, falling to 2.92 per 100,000 population in 2040 **(Figure 8)**. The number of incidence cases in almost all age groups will increase by 2040, while the incidence cases of IBD in the over-40 age group will increase substantially **(Supplementary Figure 1)**. The total number of incidence cases was the highest in the 30-34 years age group in 2019 and the highest in the 50-54 years age group in 2040, reaching 17,984 **(Supplementary Table 7)**.

**Figure 8.**
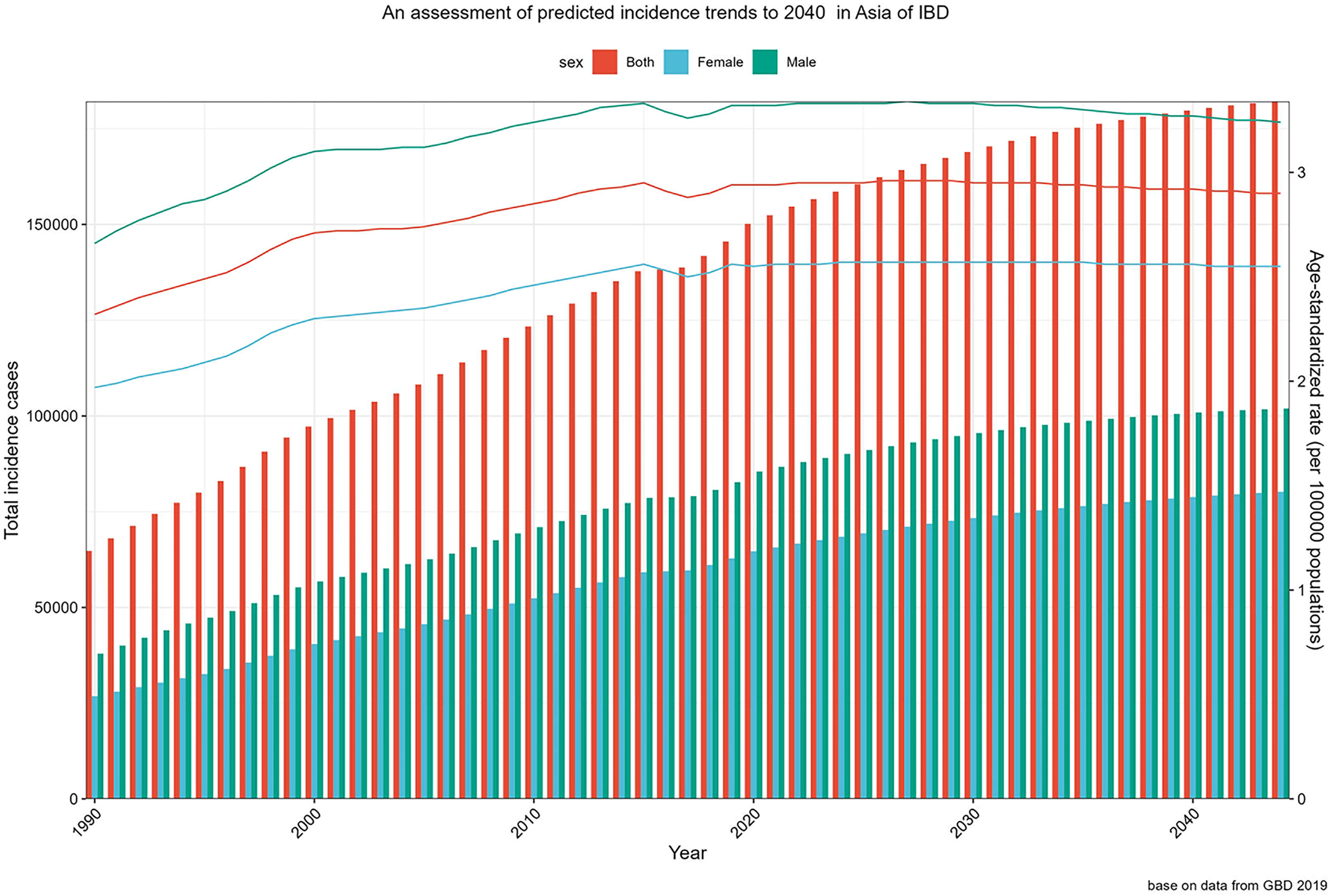
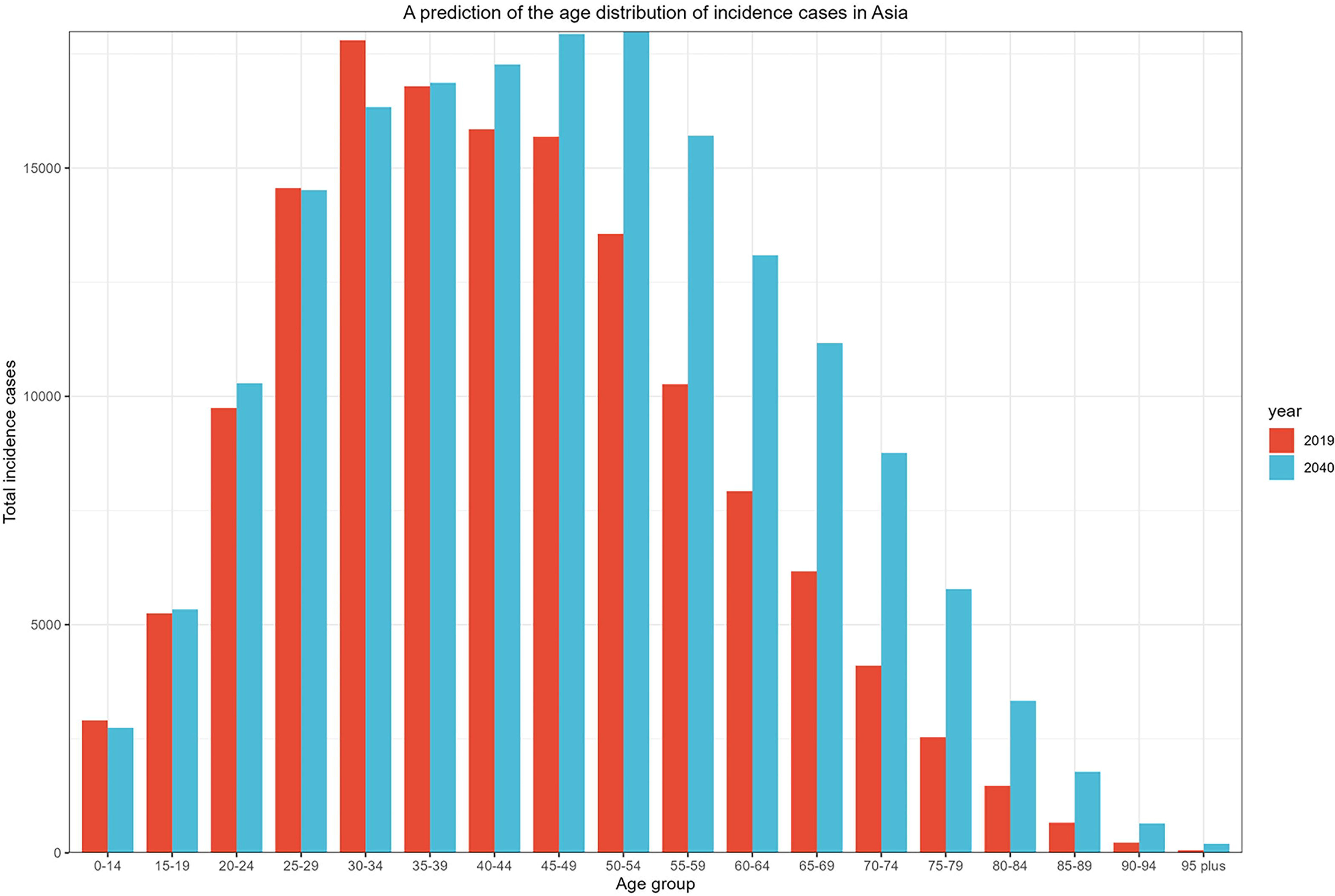
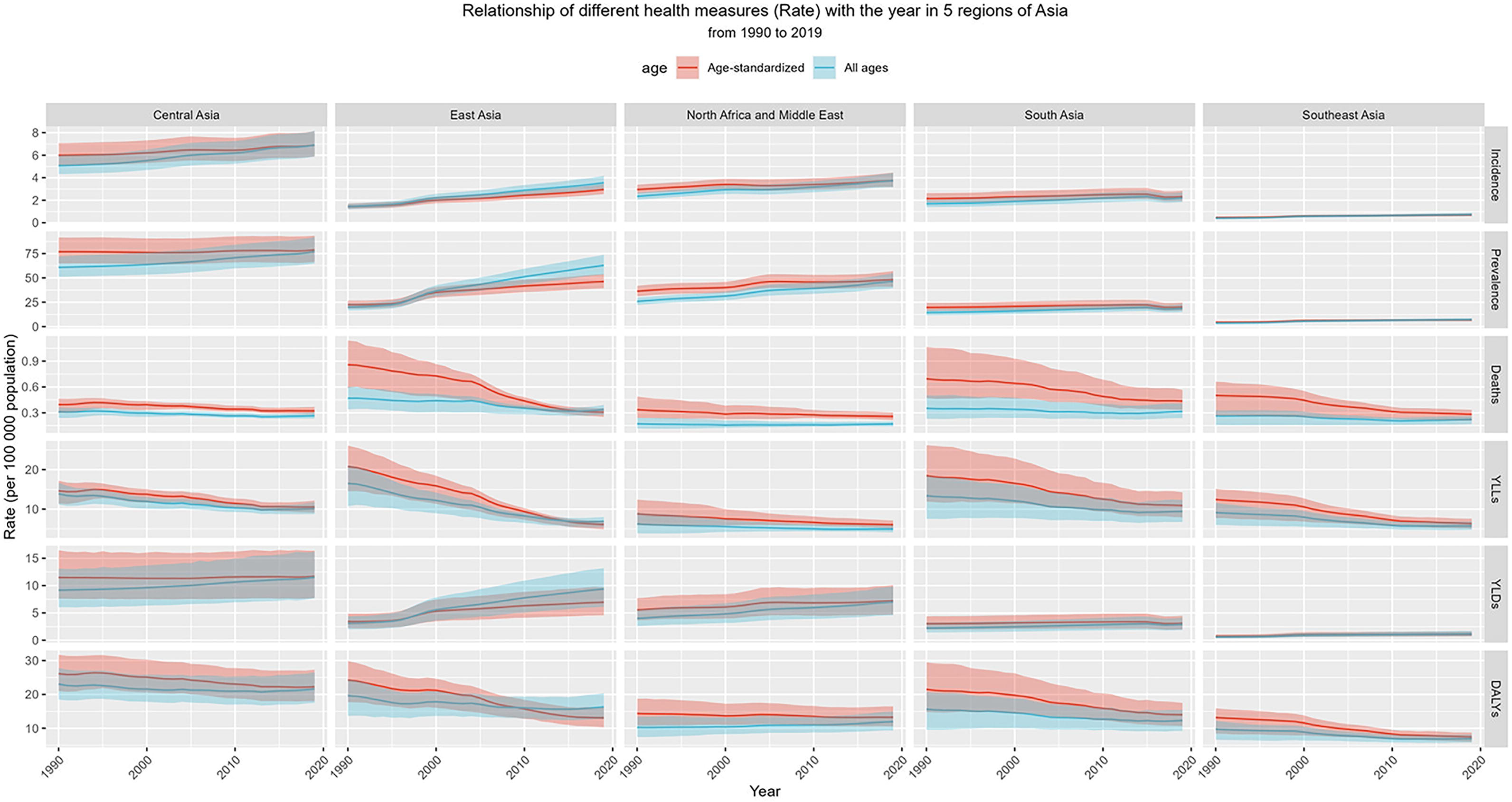
The total number and age-standardized rates of IBD incidence from 1990 to 2019 and predicted number and rates in 2040. IBD, inflammatory bowel disease.

## Discussion

IBD is a chronic and costly disease for which there is still no favorable treatment.^33, 34^ The disease has a long treatment duration, a wide age range of onset, and a high cost of standard care, which imposes a significant burden on individuals and the health care system.^6, 35, 36^ In this study, we analyzed the trends in the burden of IBD in Asia from 1990 to 2019 and projected the changes in the incidence of IBD in Asia over the next 20 years. In 2019, the number of incidence cases of IBD was 145,561 (an increase of 124.7%); the total number of prevalence cases has reached approximately 2 million (an increase of 160.92%); there were 13,957 deaths attributable to IBD (an increase of 16.03%). During this period, the total YLDs caused by IBD more than doubled to 299,663 (an increase of 158.37 %), and the total DALYs increased rapidly to 649,760. The increase in incidence, prevalence, deaths, and DALYs has led to a significant increase in the burden of IBD in Asia. The Asian IBD epidemic is heterogeneous, with the number of incidence cases maintaining elevated levels in East Asia, South Asia, and the Middle East, and the total number of prevalence cases concentrated in East Asia. Compared to several other regions, IBD-related deaths and total DALYs are significantly higher in East and South Asia, and the burden of IBD in Asia is currently concentrated in East and South Asian countries. It is worth noting that the total burden in the Middle East is relatively low, but there is a clear upward trend in incidence and DALYs. The burden of IBD in Central and Southeast Asia remains at a lower level compared with the other three regions, with no significant change in incidence, prevalence, deaths, and DALYs over 30 years **(Supplementary Figure 2)**.

In 2019, China accounted for nearly half of the total number of IBD patients in Asia, with the highest number of incidence and prevalence cases, while Japan had the highest age-standardized prevalence rate. It is noteworthy that although some countries have a low number of IBD prevalence cases, the number of incidence cases is increasing rapidly. For instance, Qatar and the United Arab Emirates experienced a staggering increase in the number of incidence cases of 862.7% and 793.6%, respectively, in 30 years, as did similar countries such as Jordan and Maldives. These countries or regions may be experiencing the early stages of an IBD epidemic but may be underestimated due to low prevalence numbers and inadequate healthcare systems.^37^ The total number of IBD-related deaths in Asia increased modestly between 1990 and 2019, but the age-standardized death rate fell by half. China, with its substantial number of prevalent cases, holds the top position in Asia regarding the total number of IBD deaths. In 2019, the number of prevalent cases in Japan was half that of China, while the actual number of IBD deaths was only one-tenth that of China. Japan, which has the highest age-standardized prevalence rate in Asia, has an extremely low age-standardized death rate, which could be attributed to its advanced healthcare system.

The rapid advancement of the economic and healthcare system has resulted in a gradual reduction in the total YLLs in Asia over time. China, Japan, India, Turkey, and the Republic of Korea were the 5 countries with the highest prevalence of IBD cases, with only India showing a significant increase in the total YLLs. Although the fatal burden of IBD is relatively low, the non-fatal burden is steadily increasing. The total YLDs in Asia have raised dramatically over time, doubling in just 30 years. The total burden of DALYs has increased significantly at the same time, with China, India, and Japan ranking as the top 3 countries with the highest burden of IBD in Asia in 2019.

The increase in the overall burden of IBD in Asia is mainly driven by population growth and aging, with incidence and DALYs continuing to rise in most countries. Our decomposition analysis revealed that the increased incidence of IBD in Asia is primarily driven by epidemiologic changes in East Asia and population growth in South and Central Asia. China, India, and Pakistan together account for more than 68% of Asia’s population, and Japan exhibits a high baseline incidence rate and IBD epidemic characteristics that differ from those of other newly industrialized countries. They exert a significant influence on the overall trend of IBD incidence in Asia. The increase in total DALYs in Asia is driven by population growth in South Asia and aging in East Asia. Despite a decline in the age-standardized mortality and total YLLs, the evolving epidemiological trends in IBD are far from offsetting the impact of population growth and aging.

The analysis shows countries with a high SDI generally have higher IBD prevalence and DALYs burden than countries with a low SDI, which is consistent with the conclusions of previous studies.^38^ Although there should be a negative correlation between age-standardized DALYs rate and healthcare quality, we conducted a frontier analysis of countries in Asia to facilitate a more comprehensive comparison of the relationship between national development status and DALYs burden. While there are leading countries at all SDI levels, it is the leading countries at low SDI levels that are most notable, such as the Lao People’s Democratic Republic, Cambodia, Yemen, etc. Despite limited in medical and economic resources, these countries continue to lead the way in addressing the burden of IBD. Meanwhile, some countries with higher SDI are also lagging, such as Japan, the Republic of Korea, and the United Arab Emirates. Countries with large gaps from the frontier should undertake work in the future to understand the factors influencing the gap between that country and the leading performers, which may be beneficial in reducing the related burden of IBD.

In this study, we predict that the overall trend of IBD incidence in Asia will continue to increase over the next 20 years. The number of IBD cases in Asia is expected to increase to 179,756 by 2040, with a higher proportion of cases occurring in older patients, which will impose a huge financial burden on every country in the region. Economic development and environmental factors have multiple influences on the increasing incidence of IBD, and some of the same environmental factors have stimulated the accelerated epidemic of IBD in countries with similar economic development. These risk factors may include high levels of urbanization, tobacco use, diets deficient in dietary fiber, rich in fat and refined sugars, as well as alterations in hygiene.^39^ In the prediction studies, we assume that the observed incidence trends will persist in the future, thus there is a degree of conservatism and uncertainty. Major public health decisions and advances in medical technology exert a profound influence on the epidemic of IBD and will require continued attention to the characteristics of this disease in the future.^40^

## Conclusions

In conclusion, population growth and aging are increasing the burden of DALYs from IBD in Asia, and this trend is projected to continue in the future. The economic, population and epidemiological characteristics of different geographical areas in Asia exhibit considerable variations.^41^ Highly developed countries in the mixed epidemic phase of IBD should aim to shorten the transition time from mixed epidemic to epidemic equilibrium in order to reduce the overall IBD burden; middle-income countries in the accelerated epidemic phase should prioritize disease prevention efforts to reduce the incidence cases of IBD; countries with lower economic levels that are currently in the ascending phase need to closely monitor the epidemic trends of IBD to minimize the future burden. Currently, the development of IBD may be at different epidemiological stages in various regions of Asia. Developing and emerging countries and regions in Asia should improve their existing healthcare delivery models and learn from the experiences of frontier countries to cope with changing population structures during IBD epidemics.

## Data availability

The data underlying this article are available in the article and in its online supplementary material.

## Declarations

The authors declare that the manuscript, including related data, figures, and tables has not been previously published and that the manuscript is not under consideration elsewhere.

## Data Availability

All data produced in the present work are contained in the manuscript

## Acknowledgments

We appreciate the invaluable contributions made by the collaborators of the Global Burden of Diseases, Injuries, and Risk Factors Study (GBD) 2019.

## Authors’ contributions

Xingchen Wang: Conceptualization, Formal Analysis, Data curation, Writing – original draft; Jiannan Xiong: Conceptualization, Data curation, Investigation; Zhao Ding: Conceptualization, Methodology, Investigation; Yueting Liu: Supervision, Writing – review & editing; Chongguang Yang: Methodology, Writing – review & editing; Pengfei Xu: Conceptualization, Funding acquisition, Resources, Supervision, Writing - review & editing.

## References

1. Lamb CA, Kennedy NA, Raine T, et al. British Society of Gastroenterology consensus guidelines on the management of inflammatory bowel disease in adults. Gut 2019;68(Suppl 3):s1–s106. doi:10.1136/gutjnl-2019-318484

2. Carter MJ, Lobo AJ, Travis SP, et al. Guidelines for the management of inflammatory bowel disease in adults. Gut 2004;53(Suppl 5):V1–16. doi:10.1136/gut.2004.043372

3. Guan Q. A Comprehensive Review and Update on the Pathogenesis of Inflammatory Bowel Disease. J Immunol Res 2019;2019:7247238. doi:10.1155/2019/7247238

4. Lautenschlager C, Schmidt C, Fischer D, Stallmach A. Drug delivery strategies in the therapy of inflammatory bowel disease. Adv Drug Deliv Rev 2014;71:58–76. doi:10.1016/j.addr.2013.10.001

5. Bernstein CN, Wajda A, Blanchard JF. The clustering of other chronic inflammatory diseases in inflammatory bowel disease: a population-based study. Gastroenterology 2005;129(3):827–36. doi:10.1053/j.gastro.2005.06.021

6. Long MD, Hutfless S, Kappelman MD, et al. Challenges in designing a national surveillance program for inflammatory bowel disease in the United States. Inflamm Bowel Dis 2014;20(2):398–415. doi:10.1097/01.MIB.0000435441.30107.8b

7. Peyrin-Biroulet L, Panes J, Sandborn WJ, et al. Defining Disease Severity in Inflammatory Bowel Diseases: Current and Future Directions. Clin Gastroenterol Hepatol 2016;14(3):348–354 e17. doi:10.1016/j.cgh.2015.06.001

8. Molodecky NA, Soon IS, Rabi DM, et al. Increasing incidence and prevalence of the inflammatory bowel diseases with time, based on systematic review. Gastroenterology 2012;142(1):46–54 e42; quiz e30. doi:10.1053/j.gastro.2011.10.001

9. Borren NZ, Van der Woude CJ, Ananthakrishnan AN. Fatigue in IBD: epidemiology, pathophysiology and management. Nat Rev Gastroenterol Hepatol 2019;16(4):247–259. doi:10.1038/s41575-018-0091-9

10. Crohn BB, Ginzburg L, Oppenheimer GD. Regional ileitis A pathologic and clinical entity. The american journal of medicine 1952(5);13:583-90. doi:10.1016/0002-9343 (52)90025-9

11. Kaplan GG. The global burden of IBD: from 2015 to 2025. Nat Rev Gastroenterol Hepatol 2015;12(12):720–7. doi:10.1038/nrgastro.2015.150

12. Aniwan S, Park SH, Loftus EV. Epidemiology, Natural History, and Risk Stratification of Crohn’s Disease. Gastroenterology Clinics of North America 2017;46(3):463–480. doi:10.1016/j.gtc.2017.05.003

13. Ng SC, Shi HY, Hamidi N, et al. Worldwide incidence and prevalence of inflammatory bowel disease in the 21st century: a systematic review of population-based studies. Lancet 2017;390:2769–2778. doi:10.1016/S0140-6736(17)32448-0

14. Kaplan GG, Windsor JW. The four epidemiological stages in the global evolution of inflammatory bowel disease. Nat Rev Gastroenterol Hepatol 2021;18(1):56–66. doi:10.1038/s41575-020-00360-x

15. Shouval DS, Rufo PA. The Role of Environmental Factors in the Pathogenesis of Inflammatory Bowel Diseases: A Review. JAMA Pediatr 2017;171(10):999–1005. doi:10.1001/jamapediatrics.2017.2571

16. Ramos GP, Papadakis KA. Mechanisms of Disease: Inflammatory Bowel Diseases. Mayo Clin Proc 2019;94(1):155–165. doi:10.1016/j.mayocp.2018.09.013

17. Nishida A, Inoue R, Inatomi O, Bamba S, Naito Y, Andoh A. Gut microbiota in the pathogenesis of inflammatory bowel disease. Clin J Gastroenterol 2018(1);11:1-10. doi:10.1007/s12328-017-0813-5

18. Ng SC, Tang W, Leong RW, et al. Environmental risk factors in inflammatory bowel disease: a population-based case-control study in Asia-Pacific. Gut 2015;64(7):1063–71. doi:10.1136/gutjnl-2014-307410

19. Mak WY, Zhao M, Ng SC, et al. The epidemiology of inflammatory bowel disease: East meets west. J Gastroenterol Hepatol 2020;35:380–389. doi:10.1111/jgh.14872

20. Ananthakrishnan AN. Epidemiology and risk factors for IBD. Nat Rev Gastroenterol Hepatol 2015;12(3):205–17. doi:10.1038/nrgastro.2015.34

21. Kaplan GG, Ng SC. Understanding and Preventing the Global Increase of Inflammatory Bowel Disease. Gastroenterology 2017;152(2):313–321 e2. doi:10.1053/j.gastro.2016.10.020

22. Ng SC, Tang W, Ching JY, et al. Incidence and phenotype of inflammatory bowel disease based on results from the Asia-pacific Crohn’s and colitis epidemiology study. Gastroenterology 2013;145(1):158–165 e2. doi:10.1053/j.gastro.2013.04.007

23. GBD 2019 Risk Factors Collaborators. Global burden of 87 risk factors in 204 countries and territories, 1990-2019: a systematic analysis for the Global Burden of Disease Study 2019. Lancet 2020;396(10258):1223-1249. doi:10.1016/S0140-6736(20)30752-2

24. GBD 2019 Diseases and Injuries Collaborators. Global burden of 369 diseases and injuries in 204 countries and territories, 1990-2019: a systematic analysis for the Global Burden of Disease Study 2019. Lancet 2020;396(10258):1204-1222. doi:10.1016/S0140-6736(20)30925-9

25. GBD 2019 Universal Health Coverage Collaborators. Measuring universal health coverage based on an index of effective coverage of health services in 204 countries and territories, 1990-2019: a systematic analysis for the Global Burden of Disease Study 2019. Lancet 2020;396(10258):1250-1284. doi:10.1016/S0140-6736(20)30750-9

26. GBD 2019 Demographics Collaborators. Global age-sex-specific fertility, mortality, healthy life expectancy (HALE), and population estimates in 204 countries and territories, 1950-2019: a comprehensive demographic analysis for the Global Burden of Disease Study 2019. Lancet 2020;396(10258):1160-1203. doi:10.1016/S0140-6736(20)30977-6

27. Hankey BF, Ries LA, Kosary CL, et al. Partitioning linear trends in age-adjusted rates. Cancer Causes Control 2000;11:31–35. doi:10.1023/a:1008953201688

28. Liu Z, Jiang Y, Yuan H, et al. The trends in incidence of primary liver cancer caused by specific etiologies: Results from the Global Burden of Disease Study 2016 and implications for liver cancer prevention. Journal of Hepatology 2019;70(4):674–683. doi:10.1016/j.jhep.2018.12.001

29. Wang R, Li Z, Liu S, et al. Global, regional and national burden of inflammatory bowel disease in 204 countries and territories from 1990 to 2019: a systematic analysis based on the Global Burden of Disease Study 2019. BMJ Open 2023;13(3):e065186. doi:10.1136/bmjopen-2022-065186

30. Moller B, Fekjaer H, Hakulinen T, et al. Prediction of cancer incidence in the Nordic countries: empirical comparison of different approaches. Stat Med 2003;22(17):2751–66. doi:10.1002/sim.1481

31. Arnold M, Park JY, Camargo MC, Lunet N, Forman D, Soerjomataram I. Is gastric cancer becoming a rare disease? A global assessment of predicted incidence trends to 2035. Gut 2020;69(5):823-829. doi:10.1136/gutjnl-2019-320234

32. Xie Y, Bowe B, Mokdad AH, et al. Analysis of the Global Burden of Disease study highlights the global, regional, and national trends of chronic kidney disease epidemiology from 1990 to 2016. Kidney Int 2018;94(3):567–581. doi:10.1016/j.kint.2018.04.011

33. Kawalec P. Indirect costs of inflammatory bowel diseases: Crohn’s disease and ulcerative colitis. A systematic review. Arch Med Sci 2016;12(2):295–302. doi:10.5114/aoms.2016.59254

34. Petryszyn PW, Witczak I. Costs in inflammatory bowel diseases. Prz Gastroenterol 2016;11(1):6–13. doi:10.5114/pg.2016.57883

35. Seyedian SS, Nokhostin F, Malamir MD. A review of the diagnosis, prevention, and treatment methods of inflammatory bowel disease. J Med Life 2019;12(2):113–122. doi:10.25122/jml-2018-0075

36. Peery AF, Crockett SD, Murphy CC, et al. Burden and Cost of Gastrointestinal, Liver, and Pancreatic Diseases in the United States: Update 2018. Gastroenterology 2019;156(1):254–272.e11. doi:10.1053/j.gastro.2018.08.063

37. Yoshida A, Ueno F, Morizane T, et al. Asian Perspectives on Diagnostic and Therapeutic Strategies in Inflammatory Bowel Disease: Report and Analysis of a Survey with Questionnaires. Digestion 2017;95(1):79–88. doi:10.1159/000453007

38. GBD 2017 Inflammatory Bowel Disease Collaborators. The global, regional, and national burden of inflammatory bowel disease in 195 countries and territories, 1990-2017: a systematic analysis for the Global Burden of Disease Study 2017. Lancet Gastroenterol Hepatol 2020;5(1):17-30. doi:10.1016/S2468-1253(19)30333-4

39. Abegunde AT, Muhammad BH, Bhatti O, Ali T. Environmental risk factors for inflammatory bowel diseases: Evidence based literature review. World Journal of Gastroenterology 2016;22(27):6296–6317. doi:10.3748/wjg.v22

40. Restall GJ, Simms AM, Walker JR, et al. Coping with Inflammatory Bowel Disease: Engaging with Information to Inform Health-Related Decision Making in Daily Life. Inflamm Bowel Dis 2017;23(8):1247–1256. doi:10.1097/MIB.0000000000001141

41. Park SH. Update on the epidemiology of inflammatory bowel disease in Asia: where are we now? Intest Res 2022;20(2):159–164. doi:10.5217/ir.2021.00115

42. https://vizhub.healthdata.org/gbd-results/. Accessed 20 July 2023.

43. http://www.kreftregisteret.no/en/Research/Projects/Nordpred/Nordpred-software/. Accessed 5 July 2023.

