## Supplementary Materials for "Inflammatory bowel disease burden in Asia from 1990 to 2019 and predictions to 2040"

**Supplementary Figures**

**Supplementary Figure 1** The prediction of the total number of IBD incidence in age groups in 2019 and 2040, both sexes.

**Supplementary Figure 2** The crude rate and age-standardized rate of different measures of IBD in 5 regions of Asia from 1990 to 2019.

**Supplementary Tables**

**Supplementary Table 1** The total number of incidence, prevalence, deaths, YLLs, YLDs, and DALYs of all ages in Asia in 1990 and 2019.

**Supplementary Table 2** The age-standardized incidence, prevalence, deaths, YLLs, YLDs, and DALYs rate (per 100,000 population) in Asia in 1990 and 2019.

**Supplementary Table 3** The total number and age-standardized rate of incidence, prevalence, and deaths in regions of Asia in 2019.

**Supplementary Table 4** The total number and age-standardized rate of YLLs, YLDs, and DALYs in regions of Asia in 2019.

**Supplementary Table 5** The decomposition analysis of IBD-related incidence and DALYs in different regions of Asia from 1990 to 2019.

**Supplementary Table 6** An assessment of predicted incidence trends to 2040 in Asia.

**Supplementary Table 7** Prediction of age-standardised incidence rate in each age group of IBD from 2020 to 2040.

**Supplementary Methods**

1. Decomposition Analysis

2. Frontier Analysis

**Supplementary Figures**

**Supplementary Figure 1 The prediction of the total number of IBD incidence in age groups in 2019 and 2040, both sexes.** IBD, inflammatory bowel disease

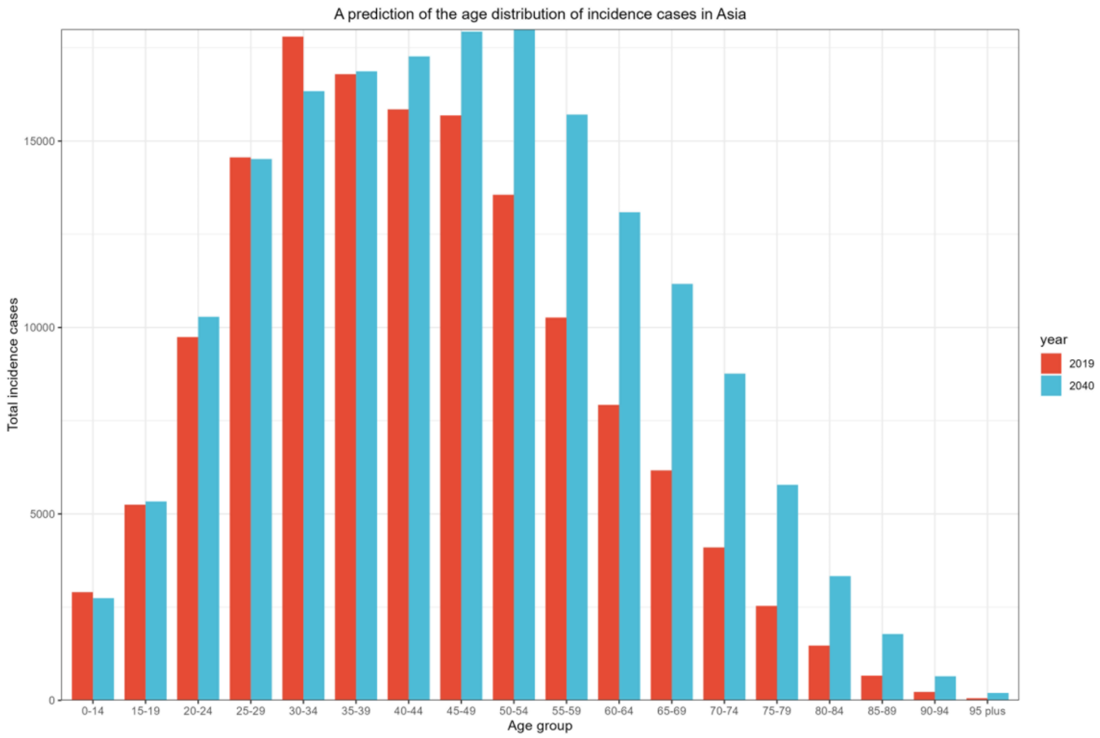

**Supplementary Figure 2 The crude rate and age-standardized rate of different measures of IBD in 5 regions of Asia from 1990 to 2019.** IBD, inflammatory bowel disease; YLLs, years of life lost; YLDs, years lived with disability; DALYs, disability-adjusted life-years.

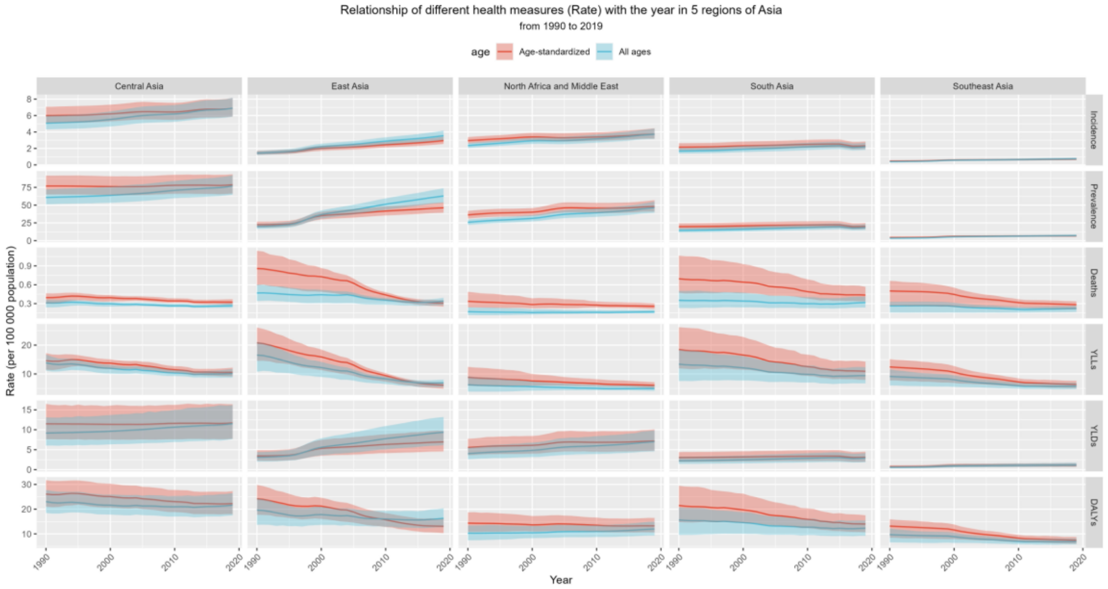

**Supplementary Table 1. The total number of incidence, prevalence, deaths, YLLs, YLDs, and DALYs of all ages in Asia in 1990 and 2019**

| | Female (95% UI) | | Male (95% UI) | | Both (95% UI) | |
|---|---|---|---|---|---|---|
| **Measure** | **1990** | **2019** | **1990** | **2019** | **1990** | **2019** |
| Incidence | 26827 (22586-31724) | 62840 (54255-73496) | 37940 (31595-45212) | 82721 (70654-97174) | 64768 (54186-76819) | 145561 (124960-170895) |
| Prevalence | 348952 (289914-412408) | 923687 (793214-1072032) | 414592 (344796-497045) | 1068534 (913134-1246870) | 763544 (634874-905119) | 1992221 (1707092-2322098) |
| Deaths | 6107 (3701-7747) | 6766 (5462-7862) | 5922 (3874-8433) | 7191 (5898-8796) | 12030 (8669-15526) | 13957 (11898-16021) |
| YLLs | 217123 (107235-279989) | 161618 (123907-191895) | 207843 (125799-275058) | 188479 (153519-235729) | 424966 (278705-534164) | 350097 (292031-409529) |
| YLDs | 54479 (35861-76966) | 141776 (94042-196499) | 61503 (40173-86698) | 157887 (104323-222128) | 115982 (75838-162919) | 299663 (198365-418635) |
| DALYs | 271603 (159080-343402) | 303394 (239582-36940) | 269346 (183761-340858) | 346366 (277839-423501) | 540949 (382320-666324) | 649760 (530395-783181) |

UI, uncertainty interval; YLLs, years of life lost; YLDs, years lived with disability; DALYs, disability-adjusted life-years

**Supplementary Table 2. The age-standardized incidence, prevalence, deaths, YLLs, YLDs, and DALYs rate (per 100,000 population) in Asia in 1990 and 2019**

| | Female (95% UI) | | Male (95% UI) | | Both (95% UI) | |
|---|---|---|---|---|---|---|
| **Measure** | **1990** | **2019** | **1990** | **2019** | **1990** | **2019** |
| Incidence | 1.97 (1.68-2.34) | 2.56 (2.21-2.98) | 2.66 (2.26-3.17) | 3.31 (2.85-3.88) | 2.32 (1.97-2.76) | 2.94 (2.53-3.44) |
| Prevalence | 27.54 (23.05-32.81) | 36.64 (31.42-42.45) | 32.10 (26.81-38.41) | 42.18 (36.08-49.20) | 29.81 (24.93-35.53) | 39.37 (33.70-45.81) |
| Deaths | 0.65 (0.40-0.89) | 0.29 (0.24-0.34) | 0.69 (0.44-1.03) | 0.36 (0.30-0.44) | 0.67 (0.46-0.90) | 0.33 (0.28-0.37) |
| YLLs | 16.45 (9.21-20.45) | 6.90 (5.27-8.17) | 16.77 (10.70-23.38) | 8.34 (6.82-10.34) | 16.60 (11.79-20.90) | 7.59 (6.32-8.88) |
| YLDs | 4.24 (2.78-5.94) | 5.64 (3.74-7.82) | 4.72 (3.12-6.65) | 6.23 (4.11-8.76) | 4.48 (2.97-6.29) | 5.93 (3.92-8.28) |
| DALYs | 20.69 (13.26-25.37) | 12.54 (9.90-15.28) | 21.49 (15.10-28.05) | 14.57 (11.80-17.67) | 21.08 (15.82-25.98) | 13.51 (11.08-16.21) |

UI, uncertainty interval; YLLs, years of life lost; YLDs, years lived with disability; DALYs, disability-adjusted life-years

**Supplementary Table 3. The total number and age-standardized rate of incidence, prevalence, and deaths in regions of Asia in 2019**

| Location | Incidence | | Prevalence | | Deaths | |
|---|---|---|---|---|---|---|
| | Incidence cases No. (95% UI) | ASIR (per 100,000) No. (95% UI) | Prevalence cases No. (95% UI) | ASPR (per 100,000) No. (95% UI) | Deaths caess No. (95% UI) | ASDR (per 100,000) No. (95% UI) |
| **Central Asia** | | | | | | |
| Armenia | 237 (200-283) | 6.64 (5.62-7.95) | 3263 (2744-3956) | 86.67 (72.86-104.56) | 11 (8-13) | 0.28 (0.21-0.34) |
| Azerbaijan | 805 (677-965) | 7.01 (5.92-8.40) | 9463 (7886-11458) | 81.93 (68.62-99.02) | 13 (10-18) | 0.16 (0.11-0.23) |
| Georgia | 275 (234-327) | 6.32 (5.38-7.44) | 4210 (3545-5060) | 87.97 (73.75-104.83) | 17 (10-22) | 0.29 (0.19-0.37) |
| Kazakhstan | 1389 (1185-1660) | 7.20 (6.14-8.56) | 15823 (13219-18930) | 81.77 (68.13-97.46) | 82 (66-100) | 0.46 (0.37-0.56) |
| Kyrgyzstan | 390 (329-460) | 6.39 (5.41-7.54) | 4283 (3592-5083) | 73.31 (61.61-87.09) | 10 (8-12) | 0.19 (0.16-0.25) |
| Mongolia | 251 (210-300) | 7.35 (6.20-8.69) | 2328 (1907-2810) | 69.72 (57.44-83.84) | 12 (9-16) | 0.45 (0.33-0.61) |
| Tajikistan | 518 (439-612) | 6.23 (5.32-7.30) | 5519 (4623-6632) | 71.19 (59.54-85.49) | 18 (13-26) | 0.30 (0.22-0.41) |
| Turkmenistan | 383 (324-451) | 7.68 (6.53-9.05) | 3806 (3155-4551) | 77.89 (64.42-92.76) | 14 (9-20) | 0.33 (0.22-0.46) |
| Uzbekistan | 520 (410-651) | 6.90 (5.90-8.20) | 5742 (4546-7126) | 75.79 (63.42-90.50) | 13 (7-22) | 0.29 (0.23-0.39) |
| **East Asia** | | | | | | |
| China | 51462 (43933-60474) | 3.01 (2.59-3.50) | 911045 (776347-1069533) | 47.06 (40.05-54.99) | 4676 (3774-5461) | 0.30 (0.24-0.35) |
| Japan | 22436 (19353-25906) | 19.65 (16.87-22.71) | 408632 (353858-468152) | 291.9 (251.75-336.57) | 452 (350-629) | 0.11 (0.09-0.16) |
| North Korea (DPRK) | 387 (326-463) | 1.29 (1.08-1.53) | 4965 (4120-5978) | 15.61 (12.96-18.71) | 108 (75-147) | 0.41 (0.28-0.59) |
| Republic of Korea | 4407 (4220-4589) | 7.33 (7.03-7.63) | 46956 (44197-49997) | 70.21 (66.03-74.79) | 234 (183-313) | 0.30 (0.23-0.39) |
| Taiwan (China) | 470 (420-526) | 1.72 (1.54-1.93) | 9064 (8139-10153) | 27.80 (24.80-31.28) | 199 (152-264) | 0.50 (0.38-0.65) |
| **Middle East** | | | | | | |
| Afghanistan | 684 (557-828) | 2.45 (2.03-2.96) | 5699 (4548-7113) | 23.69 (19.05-29.24) | 58 (31-89) | 0.43 (0.26-0.64) |
| Bahrain | 58 (48-72) | 3.21 (2.68-3.88) | 846 (688-1035) | 44.05 (36.40-53.11) | 1.2 (0.8-1.6) | 0.17 (0.13-0.22) |
| Iran (Islamic Republic of) | 3167 (2599-3894) | 3.43 (2.86-4.23) | 40493 (32952-49751) | 44.80 (36.70-55.07) | 132 (55-164) | 0.19 (0.08-0.24) |

| | | | | | | |
|---|---|---|---|---|---|---|
| Iraq | 984 (810-1191 | 2.54 (2.12-3.08) | 10029 (8221-12199) | 28.11 (23.22-34.20) | 51 (38-67) | 0.19 (0.15-0.27) |
| Jordan | 779 (649-917) | 6.91 (5.78-8.15) | 11508 (9646-13565) | 113.51 (95.78-133.87) | 13 (10-17) | 0.23 (0.18-0.30) |
| Kuwait | 195 (161-239) | 3.72 (3.09-4.54) | 2857 (2338-3483) | 54.34 (45.07-65.64) | 4 (3-5) | 0.15 (0.12-0.20) |
| Lebanon | 194 (161-238) | 3.56 (2.97-4.37) | 2494 (2037-3050) | 46.20 (37.84-56.68) | 8 (4-12) | 0.15 (0.08-0.23) |
| Oman | 143 (115-173) | 2.73 (2.27-3.27) | 1575 (1266-1924) | 33.81 (27.79-40.8) | 4 (3-5) | 0.23 (0.15-0.34) |
| Palestine | 122 (101-145) | 2.94 (2.47-3.50) | 1351 (1104-1621) | 35.42 (29.15-42.61) | 4 (3-6) | 0.20 (0.15-0.29) |
| Qatar | 125 (102-152) | 3.55 (3.00-4.28) | 1446 (1178-1746) | 44.46 (36.75-53.66) | 2 (1-3) | 0.34 (0.21-0.51) |
| Saudi Arabia | 1122 (929-1353) | 2.66 (2.24-3.18) | 13816 (11338-16751) | 34.43 (28.66-41.36) | 27 (18-38) | 0.15 (0.11-0.21) |
| Syrian Arab Republic | 449 (376-544) | 3.16 (2.63-3.83) | 4862 (3940-5871) | 33.67 (27.34-40.47) | 37 (20-52) | 0.36 (0.19-0.52) |
| Turkey | 5961 (5119-6893) | 6.44 (5.55-7.42) | 93902 (80596-107925) | 98.87 (84.96-113.45) | 329 (250-471) | 0.40 (0.30-0.56) |
| United Arab Emirates | 520 (410-651) | 4.12 (3.40-5.01) | 5742 (4546-7126) | 42.72 (35.58-51.97) | 13 (7-22) | 0.24 (0.15-0.43) |
| Yemen | 647 (527-793) | 2.54 (2.10-3.09) | 6284 (5044-7734) | 28.03 (22.65-34.94) | 44 (30-68) | 0.33 (0.22-0.52) |
| **South Asia** | | | | | | |
| Cambodia | 79 (64-96) | 0.49 (0.40-0.59) | 595 (467-730) | 3.77 (3.01-4.61) | 38 (26-48) | 0.35 (0.23-0.45) |
| Bangladesh | 3404 (2788-4136) | 2.17 (1.78-2.65) | 33018 (26285-41258) | 21.62 (17.31-27) | 713 (431-1068) | 0.56 (0.35-0.83) |
| Bhutan | 14 (12-17) | 1.86 (1.55-2.29) | 158 (128-197) | 22.23 (18.11-27.62) | 3 (2-7) | 0.55 (0.34-1.23) |
| India | 31775 (26433-38972) | 2.34 (1.95-2.86) | 270719 (219873-332264) | 20.34 (16.57-24.97) | 4214 (2950-5531) | 0.40 (0.29-0.53) |
| Nepal | 624 (515-764) | 2.24 (1.85-2.76) | 5036 (4049-6194) | 18.72 (15.04-23.18) | 115 (71-190) | 0.59 (0.36-0.98) |
| Pakistan | 3888 (3193-4838) | 2.30 (1.91-2.86) | 26815 (21768-33849) | 16.77 (13.78-21.25) | 657 (447-993) | 0.62 (0.41-0.99) |
| **Southeast Asia** | | | | | | |
| Brunei Darussalam | 26 (21-31) | 5.09 (4.17-6.09) | 295 (237-361) | 57.28 (46.33-70.04) | 2.6 (2.1-3.2) | 1.48 (0.97-1.89) |
| Indonesia | 1504 (1248-1808) | 0.55 (0.46-0.66) | 11410 (9158-13704) | 4.16 (3.37-5.00) | 913 (582-1179) | 0.55 (0.33-0.70) |
| Lao PDR | 35 (29-43) | 0.51 (0.42-0.63) | 241 (191-300) | 3.67 (2.95-4.55) | 11 (6-17) | 0.26 (0.14-0.41) |
| Malaysia | 251 (223-283) | 0.76 (0.67-0.85) | 2165 (1879-2498) | 6.56 (5.71-7.56) | 41 (30-56) | 0.17 (0.12-0.23) |
| Maldives | 2.8 (2.3-3.5) | 0.51 (0.43-0.61) | 27 (21-33) | 4.94 (3.96-5.96) | 0.9 (0.7-1.1) | 0.30 (0.23-0.40) |
| Myanmar | 306 (253-378) | 0.55 (0.46-0.68) | 2062 (1656-2558) | 3.71 (3.00-4.59) | 59 (44-82) | 0.13 (0.10-0.17) |

| | | | | | | |
|---|---|---|---|---|---|---|
| Philippines | 657 (541-799) | 0.63 (0.52-0.76) | 5073 (4044-6166) | 4.96 (4.02-6.00) | 117 (95-155) | 0.16 (0.13-0.20) |
| Singapore | 215 (178-264) | 3.21 (2.68-3.90) | 3179 (2593-3962) | 41.88 (34.28-51.91) | 5 (4-8) | 0.07 (0.06-0.11) |
| Sri Lanka | 268 (239-301) | 1.15 (1.02-1.30) | 2295 (2003-2637) | 9.51 (8.29-10.90) | 21 (15-30) | 0.10 (0.07-0.13) |
| Thailand | 373 (309-455) | 0.46 (0.38-0.56) | 3808 (3078-4630) | 4.28 (3.44-5.22) | 111 (80-149) | 0.12 (0.08-0.15) |
| Timor-Leste | 6 (5-7) | 0.51 (0.42-0.62) | 41 (33-50) | 3.90 (3.14-4.72) | 3 (2-5) | 0.36 (0.20-0.61) |
| Viet Nam | 1482 (1201-1836) | 1.37 (1.11-1.70) | 20278 (16050-25063) | 18.35 (14.68-22.55) | 177 (119-247) | 0.22 (0.15-0.32) |

UI, uncertainty interval; ASIR, age-standardized incidence rate; ASPR, age-standardized prevalence rate; ASDR, age-standardized deaths rate

**Supplementary Table 4. The total number and age-standardized rate of YLLs, YLDs, and DALYs in regions of Asia in 2019**

| Location | YLLs | | YLDs | | DALYs | |
|---|---|---|---|---|---|---|
| | Total YLLs No. (95% UI) | ASR (per 100,000) No. (95% UI) | Total YLDs No. (95% UI) | ASR (per 100,000) No. (95% UI) | Total DALYs No. (95% UI) | ASR (per 100,000) No. (95% UI) |
| **Central Asia** | | | | | | |
| Armenia | 285 (218-347) | 7.67 (6.05-9.41) | 479 (314-696) | 12.83 (8.32-18.60) | 764 (593-979) | 20.50 (15.81-26.35) |
| Azerbaijan | 418 (307-581) | 4.03 (3.03-5.50) | 1411 (918-2033) | 12.17 (7.97-17.50) | 1829 (1289-2470) | 16.20 (11.65-21.65) |
| Georgia | 378 (262-483) | 7.25 (5.31-9.12) | 612 (399-882) | 13.00 (8.37-18.79) | 991 (741-1269) | 20.25 (15.35-26.28) |
| Kazakhstan | 2819 (2304-3518) | 14.83 (12.13-18.29) | 2353 (1532-3415) | 12.13 (7.91-17.57) | 5172 (4164-6466) | 26.97 (21.78-33.61) |
| Kyrgyzstan | 386 (320-464) | 6.43 (5.38-7.82) | 645 (418-931) | 10.95 (7.11-15.71) | 1031 (789-1325) | 17.37 (13.34-22.37) |
| Mongolia | 462 (325-640) | 14.29 (10.17-19.70) | 352 (229-497) | 10.45 (6.81-14.72) | 814 (620-1028) | 24.75 (19.13-31.11) |
| Tajikistan | 808 (547-1202) | 9.95 (7.01-14.29) | 838 (531-1193) | 10.65 (6.88-15.06) | 1646 (1219-2185) | 20.60 (15.55-26.90) |
| Turkmenistan | 547 (344-794) | 11.25 (7.2-16.15) | 573 (380-822) | 11.65 (7.75-16.70) | 1120 (805-1438) | 22.90 (16.63-29.27) |
| Uzbekistan | 3355 (2682-4163) | 10.62 (8.67-13.09) | 3514 (2259-4944) | 11.28 (7.35-15.76) | 6869 (5391-8500) | 21.90 (17.37-27.16) |
| **East Asia** | | | | | | |
| China | 96558 (75917-112804) | 6.02 (4.78-6.95) | 135906 (89067-191529) | 7.07 (4.65-9.86) | 232464 (179903-291090) | 13.10 (10.29-16.31) |
| Japan | 6680 (5569-9187) | 2.48 (2.16-3.50) | 60744 (40135-83792) | 44.1 (29.09-61.15) | 67424 (46877-91495) | 46.58 (31.41-63.54) |
| North Korea (DPRK) | 2364 (1601-3219) | 8.58 (5.85-11.80) | 797 (514-1141) | 2.52 (1.62-3.60) | 3161 (2323-4117) | 11.10 (8.17-14.58) |
| Republic of Korea | 2364 (1601-3219) | 4.32 (3.46-6.56) | 797 (514-1141) | 10.68 (7.15-14.76) | 10451 (7829-13343) | 15.00 (11.13-19.33) |
| Taiwan (China) | 2930 (2248-3889) | 8.04 (6.21-10.65) | 1408 (916-1972) | 4.36 (2.85-6.13) | 4338 (3499-5416) | 12.40 (10.06-15.44) |
| **Middle East** | | | | | | |
| Afghanistan | 2125 (1018-3729) | 10.94 (5.95-16.65) | 892 (565-1318) | 3.65 (2.30-5.35) | 3017 (1879-4660) | 14.58 (9.58-20.47) |
| Bahrain | 40 (27-55) | 3.52 (2.55-4.70) | 127 (80-184) | 6.54 (4.15-9.44) | 167 (116-229) | 10.05 (7.40-13.28) |
| Iran (Islamic Republic of) | 3564 (1516-4438) | 4.53 (1.93-5.65) | 1565 (992-2275) | 6.49 (4.20-9.30) | 9481 (6679-12439) | 11.03 (7.60-14.22) |

| | | | | | | |
|---|---|---|---|---|---|---|
| Iraq | 2153 (1482-2983) | 6.09 (4.44-8.04) | 1565 (992-2275) | 4.33 (2.73-6.26) | 3719 (2808-4931) | 10.42 (8.08-13.32) |
| Jordan | 427 (324-558) | 5.19 (4.00-6.65) | 1728 (1113-2514) | 16.81 (11.07-24.25) | 2155 (1536-2953) | 22.01 (16.06-29.65) |
| Kuwait | 124 (99-157) | 3.62 (2.87-4.65) | 435 (274-630) | 8.11 (5.19-11.74) | 558 (391-755) | 11.73 (8.74-15.53) |
| Lebanon | 177 (94-272) | 3.37 (1.80-5.16) | 368 (240-523) | 6.81 (4.45-9.72) | 545 (394-737) | 10.18 (7.34-13.75) |
| Oman | 133 (95-186) | 4.86 (3.53-6.65) | 242 (151-351) | 5.10 (3.25-7.36) | 375 (271-496) | 9.96 (7.66-12.72) |
| Palestine | 113 (82-189) | 4.11 (3.06-6.47) | 208 (128-305) | 5.36 (3.42-7.79) | 320 (235-443) | 9.47 (7.18-12.75) |
| Qatar | 78 (47-114) | 6.52 (3.91-9.54) | 219 (138-327) | 6.55 (4.21-9.58) | 296 (205-412) | 13.06 (9.28-17.59) |
| Saudi Arabia | 1039 (625-1534) | 3.65 (2.47-4.98) | 2142 (1319-3179) | 5.25 (3.33-7.66) | 3181 (2284-4376) | 8.90 (6.65-11.75) |
| Syrian Arab Republic | 914 (560-1293) | 7.62 (4.49-10.65) | 739 (466-1104) | 5.13 (3.26-7.68) | 1653 (1195-2195) | 12.75 (9.00-16.93) |
| Turkey | 8492 (6513-11659) | 10.33 (7.95-13.81) | 13996 (9188-19772) | 14.73 (9.72-20.87) | 22488 (17253-28896) | 25.05 (19.42-32.24) |
| United Arab Emirates | 534 (303-913) | 6.04 (3.76-10.69) | 867 (532-1289) | 6.36 (4.02-9.32) | 1401 (971-1965) | 12.40 (8.88-17.45) |
| Yemen | 1438 (938-2152) | 8.08 (5.39-12.24) | 985 (625-1443) | 4.32 (2.75-6.33) | 2423 (1790-3248) | 12.40 (9.18-16.77) |
| **South Asia** | | | | | | |
| Cambodia | 1123 (798-1438) | 8.36 (5.84-10.65) | 99 (61-149) | 0.62 (0.39-0.93) | 1222 (890-1546) | 8.98 (6.42-11.33) |
| Bangladesh | 21249 (12680-31860) | 14.77 (8.90-22.04) | 5105 (3208-7461) | 3.33 (2.08-4.88) | 26354 (17363-37248) | 18.10 (12.11-25.27) |
| Bhutan | 77 (44-176) | 12.42 (7.37-28.66) | 24 (15-36) | 3.40 (2.13-4.93) | 102 (67-202) | 15.82 (10.39-31.52) |
| India | 122827 (81271-163422) | 9.93 (6.68-13.18) | 41590 (27306-60763) | 3.11 (2.03-4.56) | 164416 (117527-208276) | 13.04 (9.44-16.5) |
| Nepal | 3029 (1902-4983) | 12.73 (7.88-21.03) | 790 (502-1165) | 2.92 (1.85-4.32) | 3819 (2683-5781) | 15.65 (10.93-23.88) |
| Pakistan | 23051 (14815-34284) | 15.17 (10.24-22.68) | 4235 (2713-6139) | 2.62 (1.70-3.81) | 27286 (18727-38379) | 17.80 (12.91-25.37) |
| **Southeast Asia** | | | | | | |
| Brunei Darussalam | 69 (53-98) | 24.07 (19.52-29.89) | 45 (28-67) | 8.69 (5.47-12.89) | 114 (89-147) | 32.76 (26.62-39.71) |
| Indonesia | 23069 (16619-30317) | 10.98 (7.55-14.24) | 1874 (1170-2767) | 0.68 (0.43-1.00) | 24943 (18268-32252) | 11.66 (8.16-14.98) |
| Lao PDR | 347 (198-574) | 6.36 (3.59-10.57) | 40 (25-60) | 0.61 (0.38-0.91) | 387 (234-610) | 6.97 (4.13-11.24) |
| Malaysia | 1079 (782-1429) | 3.85 (2.79-5.08) | 356 (227-507) | 1.08 (0.70-1.53) | 1434 (1090-1814) | 4.92 (3.71-6.26) |
| Maldives | 25 (19-33) | 6.39 (4.97-8.46) | 4 (3-7) | 0.80 (0.51-1.18) | 29 (23-38) | 7.20 (5.69-9.23) |
| Myanmar | 1916 (1356-2910) | 3.73 (2.71-5.48) | 341 (212-517) | 0.61 (0.38-0.92) | 2258 (1680-3215) | 4.34 (3.28-6.04) |

| | | | | | | |
|---|---|---|---|---|---|---|
| Philippines | 3721 (3019-5076) | 4.06 (3.29-5.49) | 835 (516-1254) | 0.81 (0.51-1.21) | 4556 (3797-5981) | 4.87 (4.06-6.26) |
| Singapore | 126 (101-184) | 1.71 (1.38-2.50) | 487 (305-710) | 6.45 (4.07-9.52) | 612 (427-838) | 8.16 (5.69-11.25) |
| Sri Lanka | 522 (366-736) | 2.22 (1.57-3.10) | 373 (240-519) | 1.55 (1.00-2.16) | 895 (684-1147) | 3.77 (2.86-4.79) |
| Thailand | 2645 (1908-3567) | 2.93 (2.13-3.94) | 626 (393-936) | 0.71 (0.44-1.05) | 3272 (2453-4259) | 3.64 (2.76-4.68) |
| Timor-Leste | 80 (42-132) | 8.50 (4.60-14.26) | 7 (4-10) | 0.64 (0.41-0.94) | 86 (48-138) | 9.14 (5.17-14.89) |
| Viet Nam | 4134 (2835-5638) | 4.62 (3.17-6.34) | 3285 (2015-5039) | 2.98 (1.84-4.56) | 7419 (5510-9604) | 7.60 (5.65-9.80) |

UI, uncertainty interval; ASR, age-standardized rate (per 100 000 population); YLLs, years of life lost; YLDs, years lived with disability; DALYs, disability-adjusted life-years

**Supplementary Table 5. The decomposition analysis of IBD-related incidence and DALYs in different regions of Asia from 1990 to 2019**

| | Incidence | | | | | | | DALYs | | | | | | |
|---|---|---|---|---|---|---|---|---|---|---|---|---|---|---|
| **Location** | **overall** | **a_effect** | **p_effect** | **e_effect** | **a_percent** | **p_percent** | **e_percent** | **overall** | **a_effect** | **p_effect** | **e_effect** | **a_percent** | **p_percent** | **e_percent** |
| **Asia** | 80793 | 20648 | 35496 | 24648 | 25.56 | 43.94 | 30.51 | 108812 | 151295 | 214462 | -256946 | 139.04 | 197.09 | -236.14 |
| **Central Asia** | 2947 | 830 | 1447 | 670 | 28.16 | 49.10 | 22.74 | 4285 | 2289 | 5400 | -3404 | 53.42 | 126.02 | -79.44 |
| **East Asia** | 34751 | 6214 | 6049 | 22489 | 17.88 | 17.41 | 64.71 | -268 | 78512 | 44688 | -123469 | -29293.5 | -16673.6 | 46067.05 |
| **Middle East*** | 14616 | 1204 | 9222 | 4189 | 8.24 | 63.10 | 28.66 | 37317 | 11043 | 29409 | -3135 | 29.59 | 78.81 | -8.40 |
| **South Asia** | 21265 | 3385 | 14565 | 3315 | 15.92 | 68.49 | 15.59 | 50530 | 31086 | 100169 | -80725 | 61.52 | 198.24 | -159.76 |
| **Southeast Asia** | 3098 | 584 | 1168 | 1347 | 18.83 | 37.70 | 43.47 | 1370 | 11914 | 17501 | -28046 | 869.87 | 1277.85 | -2047.72 |

IBD, inflammatory bowel disease; DALYs, disability-adjusted life-years; a_effect, aging effect; p_effect, population effect; e_effect, epidemiological changes effect; a_percent, aging percent; p_percent, population percent; e_percent, epidemiological changes percent. *Middle East, North Africa and Middle East, including data for the North African region

**Supplementary Table 6. An assessment of predicted incidence trends to 2040 in Asia**

| Year | Total incidence cases | | | ASR (per 100,000) | | |
|---|---|---|---|---|---|---|
| | Female | Male | Both | Female | Male | Both |
| 2019 | 62840 | 82721 | 145561 | 2.56 | 3.32 | 2.94 |
| 2020 | 64661 | 85499 | 150160 | 2.55 | 3.32 | 2.94 |
| 2021 | 65676 | 86744 | 152421 | 2.56 | 3.32 | 2.94 |
| 2022 | 66677 | 87975 | 154652 | 2.56 | 3.33 | 2.95 |
| 2023 | 67591 | 89041 | 156632 | 2.56 | 3.33 | 2.95 |
| 2024 | 68492 | 90088 | 158580 | 2.57 | 3.33 | 2.95 |
| 2025 | 69375 | 91115 | 160490 | 2.57 | 3.33 | 2.95 |
| 2026 | 70242 | 92123 | 162364 | 2.57 | 3.33 | 2.96 |
| 2027 | 71094 | 93115 | 164210 | 2.57 | 3.34 | 2.96 |
| 2028 | 71862 | 93953 | 165816 | 2.57 | 3.33 | 2.96 |
| 2029 | 72613 | 94771 | 167384 | 2.57 | 3.33 | 2.96 |
| 2030 | 73345 | 95566 | 168911 | 2.57 | 3.33 | 2.95 |
| 2031 | 74058 | 96336 | 170394 | 2.57 | 3.32 | 2.95 |
| 2032 | 74756 | 97088 | 171844 | 2.57 | 3.32 | 2.95 |
| 2033 | 75360 | 97675 | 173034 | 2.57 | 3.31 | 2.95 |
| 2034 | 75941 | 98232 | 174173 | 2.57 | 3.31 | 2.94 |
| 2035 | 76499 | 98759 | 175258 | 2.57 | 3.30 | 2.94 |
| 2036 | 77032 | 99254 | 176286 | 2.56 | 3.29 | 2.93 |
| 2037 | 77545 | 99722 | 177267 | 2.56 | 3.28 | 2.93 |
| 2038 | 78007 | 100153 | 178160 | 2.56 | 3.28 | 2.92 |
| 2039 | 78442 | 100549 | 178990 | 2.56 | 3.27 | 2.92 |
| 2040 | 78849 | 100907 | 179756 | 2.56 | 3.27 | 2.92 |
| 2041 | 79228 | 101227 | 180455 | 2.55 | 3.26 | 2.91 |
| 2042 | 79584 | 101513 | 181096 | 2.55 | 3.25 | 2.91 |
| 2043 | 79904 | 101747 | 181651 | 2.55 | 3.25 | 2.90 |
| 2044 | 80195 | 101942 | 182138 | 2.55 | 3.24 | 2.90 |

ASR, age-standardized rate (per 100,000 population)

**Supplementary Table 7. Prediction of age-standardised incidence rate in each age group of IBD from 2020 to 2040**

| Age group | 2020 | | 2030 | | 2040 | | 2044 | |
|---|---|---|---|---|---|---|---|---|
| | Female | Male | Female | Male | Female | Male | Female | Male |
| 1 to 4 | 0.25 | 0.30 | 0.25 | 0.32 | 0.26 | 0.33 | 0.26 | 0.34 |
| 5 to 9 | 1.25 | 1.70 | 1.27 | 1.75 | 1.28 | 1.78 | 1.28 | 1.79 |
| 10 to 14 | 2.27 | 3.25 | 2.25 | 3.23 | 2.26 | 3.28 | 2.27 | 3.30 |
| 15 to 19 | 3.29 | 4.63 | 3.18 | 4.49 | 3.18 | 4.50 | 3.18 | 4.52 |
| 20 to 24 | 4.02 | 5.36 | 3.90 | 5.19 | 3.81 | 5.03 | 3.81 | 5.06 |
| 25 to 29 | 4.42 | 5.65 | 4.33 | 5.43 | 4.15 | 5.16 | 4.10 | 5.06 |
| 30 to 34 | 4.61 | 5.78 | 4.54 | 5.54 | 4.37 | 5.25 | 4.28 | 5.12 |
| 35 to 39 | 4.60 | 5.74 | 4.64 | 5.65 | 4.50 | 5.32 | 4.44 | 5.22 |
| 40 to 44 | 4.34 | 5.41 | 4.57 | 5.66 | 4.46 | 5.29 | 4.41 | 5.14 |
| 45 to 49 | 4.02 | 5.04 | 4.35 | 5.44 | 4.34 | 5.25 | 4.29 | 5.09 |
| 50 to 54 | 3.76 | 4.72 | 3.99 | 5.03 | 4.18 | 5.16 | 4.16 | 5.05 |
| 55 to 59 | 3.47 | 4.41 | 3.63 | 4.63 | 3.91 | 4.91 | 4.01 | 5.00 |
| 60 to 64 | 3.28 | 4.20 | 3.38 | 4.35 | 3.58 | 4.56 | 3.72 | 4.70 |
| 65 to 69 | 3.08 | 3.98 | 3.18 | 4.16 | 3.31 | 4.29 | 3.34 | 4.32 |
| 70 to 74 | 2.89 | 3.79 | 3.06 | 4.09 | 3.13 | 4.15 | 3.22 | 4.23 |
| 75 to 79 | 2.74 | 3.73 | 2.91 | 4.09 | 2.99 | 4.19 | 2.97 | 4.16 |
| 80 to 84 | 2.76 | 4.05 | 2.89 | 4.40 | 3.04 | 4.65 | 3.09 | 4.71 |
| 85 to 89 | 2.87 | 4.24 | 2.94 | 4.57 | 3.10 | 4.88 | 3.15 | 4.98 |
| 90 to 94 | 0.25 | 0.30 | 0.25 | 0.32 | 0.26 | 0.33 | 0.26 | 0.34 |
| 95 plus | 1.25 | 1.70 | 1.27 | 1.75 | 1.28 | 1.78 | 1.28 | 1.79 |

Age standardized incidence rate (per 100 000 population)

**Supplementary Methods**

**1. Decomposition Analysis**

The incidence and DALYs of IBD were decomposed according to population age group, population growth, and epidemiological changes using the decomposition method of Das Gupta. In the case of DALYs, the number of DALYs in each location was calculated by the following formula:

$$\boldsymbol{DALY}_{\mathrm{ay,py,ey}} = \sum_{i=1}^{20} (\boldsymbol{a}_{\mathrm{i,y}} * \boldsymbol{p}_{\mathrm{y}} * \boldsymbol{e}_{\mathrm{i,y}})$$

**DALY** $_{\mathrm{ay,\,py,\,ey}}$ represent DALYs based on factors such as age group, population, and DALYs rate for a particular year. **a** $_{\mathrm{i,\,y}}$ represents the proportion of the population in the **i** age category out of 20 age categories in a given year **y**; **p** $_{\mathrm{y}}$ represents the total population of y in a given year; **e** $_{\mathrm{i,\,y}}$ indicates the DALYs rate annuity rate for the **i** age group in year **y**. The contribution of each factor to the change in DALYs from 1990 to 2019 was defined as the effect of one factor changing while the others remained constant. The calculation formula is as follows:

$$\begin{aligned}
&[(\boldsymbol{DALY}_{\mathrm{a2019,p1990,e1990}} + \boldsymbol{DALY}_{\mathrm{a2019,p2019,e2019}})/3 \\
&+ (\boldsymbol{DALY}_{\mathrm{a2019,p1990,e2019}} + \boldsymbol{DALY}_{\mathrm{a2019,p2019,e1990}})/6] \\
&- [(\boldsymbol{DALY}_{\mathrm{a1990,p2019,e2019}} + \boldsymbol{DALY}_{\mathrm{a1990,p1990,e1990}})/3 \\
&+ (\boldsymbol{DALY}_{\mathrm{a1990,p2019,e1990}} + \boldsymbol{DALY}_{\mathrm{a1990,p1990,e2019}})/6]
\end{aligned}$$

### 2. Frontier Analysis

To assess the relationship between the burden of IBD and socio-demographic development, we applied frontier analysis as a quantitative method to determine the lowest achievable age-standardized DALYs rate based on the developmental status of SDI measurements. The DALYs frontier pinpoints determine the minimum DALYs achievable for each country or region given its SDI. The distance from the frontier is denoted by the effective difference; large effective differences from the frontier indicate that there may be unrealized gains or opportunities for improvement (such as a reduction in DALYs for IBD) depending on where the country or region is on the development spectrum. A data envelope analysis was performed using the free disposal hull method, which utilizes data from 1990-2019 and generates frontier for age-adjusted IBD DALYs via the SDI index, allowing for the delineation of nonlinear frontiers. In order to account for uncertainty, the analysis used 500 bootstrapped samples of the data, randomly sampling with replacements from all countries and regions of Asia in all years. Mean IBD DALYs were calculated for the SDI values of each bootstrapped sample. Then the smooth frontier is obtained by regression of LOESS with a local polynomial degree of 1 and a span of 0.2. To exclude the effect of outliers, super-efficient countries or locations are excluded from the frontier generation. To understand the relationship between 2019 age-standardized IBD DALYs rate and the vis-à-vis frontier, we calculated the effective difference (absolute distance from the frontier) using the 2019 SDI and age-standardized IBD DALYs rate data points for each country or location. Countries or locations with lower DALYs than the frontiers were assigned a zero distance.
